# Cost-effectiveness of PCV20 to Prevent Pneumococcal Disease in the Pediatric Population - A German Societal Perspective Analysis

**DOI:** 10.1101/2024.03.14.24304296

**Authors:** An Ta, Felicitas Kühne, Maren Laurenz, Christof von Eiff, Sophie Warren, Johnna Perdrizet

## Abstract

**Background:** The 13-valent pneumococcal conjugate vaccine (PCV13) has been included by Germany’s Standing Committee on Vaccinations for infants since 2009, resulting in major reductions in pneumococcal disease (PD). Higher-valent vaccines may further reduce PD burden. This cost-effectiveness analysis compared PCV20 under 3+1 schedule with PCV15 and PCV13, both under 2+1 schedule, in Germany’s pediatric population.

**Methods:** A Markov model with annual cycles over a 10-year time horizon was adapted to simulate the clinical and economic consequences to the German population and compare pediatric vaccination with PCV20 to lower-valent PCVs. The model used PCV13 clinical effectiveness and impact studies as well as PCV7 efficacy studies for vaccine direct and indirect effect estimates. Epidemiologic, utility, and medical cost inputs were obtained from published sources. Benefits and costs were discounted at 3% from a German societal perspective. Outcomes included PD cases, deaths, costs, quality-adjusted life years (QALYs), and incremental cost-effectiveness ratios (ICERs).

**Results:** In the base case, PCV20 provided greater health benefits than PCV13, averting more cases of invasive pneumococcal disease (IPD; 15,301), hospitalized and non-hospitalized pneumonia (460,197 and 472,365, respectively), otitis media (531,634), and 59,265 deaths over 10 years. This resulted in 904,854 additional QALYs and a total cost-saving of €2,393,263,611, making PCV20 a dominant strategy compared with PCV13. Compared to PCV15, PCV20 was estimated to avert an additional 11,334 IPD, 704,948 pneumonia, and 441,643 otitis media cases, as well as 41,596 deaths. PCV20 was associated with a higher QALY gain and lower cost (i.e., dominance) compared with PCV15. The robustness of the results was confirmed through scenario analyses as well as deterministic and probabilistic sensitivity analyses.

**Conclusion:** PCV20 3+1 dominated both PCV13 2+1 and PCV15 2+1 over the model time horizon. Replacing lower-valent PCVs with PCV20 would result in greater clinical and economic benefits, given PCV20’s broader serotype coverage.

**Key Summary Points:**

- *Streptococcus pneumoniae* is the leading cause of bacterial pneumonia and global mortality in children.
- Pneumococcal conjugate vaccines (PCVs) elicit robust and durable immune responses in both pediatric and adult populations.
- This study examined the cost-effectiveness of PCV20 under a 3+1 schedule in Germany’s pediatric population compared with PCV13 and a secondary comparator (PCV15), both under a 2+1 schedule.
- PCV20 was estimated to prevent more pneumococcal disease cases and deaths versus PCV13 and PCV15, as well as providing greater quality-adjusted life years and cost savings (i.e., dominant strategy) over 10 years.
- Implementation of PCV20 under a 3+1 schedule into the German pediatric immunization program would result in greater clinical and economic benefits versus PCV13 and PCV15, both under a 2+1 schedule.

**Plain language summary:** Pneumococcal diseases (e.g., ear infections, pneumonia, bloodstream infections) are among the leading causes of illness and death in children worldwide. The pneumococcal conjugate vaccine (PCV) protects against pneumococcal diseases and has significantly reduced the number of newly diagnosed cases. Higher-valent vaccines (which provide coverage for a greater number of disease-causing serotypes) have recently received EC approval for use in adults and EC approval for use in infants is expected soon. This study examined costs and health benefits associated with the 20-valent PCV (PCV20) under a 3+1 (i.e., three primary doses and one booster dose) schedule in Germany’s childhood vaccination program compared with 13-valent PCV (PCV13) and the 15-valent PCV (PCV15), both under a 2+1 (two primary doses, one booster) schedule. PCV20 was estimated to result in greater health benefits from avoiding more cases in pneumococcal diseases and lower costs compared with both PCV13 and PCV15. PCV20, therefore, is considered the best option among the three vaccines for children in Germany.

## Introduction

*Streptococcus pneumoniae* is the leading cause of bacterial pneumonia and global mortality in children [1–4]. In 2016, *S. pneumoniae* has been estimated to account for approximately 197 million cases of pneumonia and 1.1 million deaths[5]. This encapsulated bacterium is the major cause of pneumococcal diseases ranging from otitis media (OM) and pneumonia to life-threatening invasive pneumococcal diseases (IPD), including sepsis and meningitis.

Pneumococcal conjugate vaccines (PCV) elicit robust and durable immune responses in both pediatric and adult populations [3]. They have noticeably reduced IPD incidence across all age groups due to indirect effects (i.e., herd effects or the effect on the unvaccinated population) [6, 7]. The 7-valent PCV (PCV7) was first approved in Europe in 2001 [8], and was recommended for high-risk children in July 2001 by Germany’s Standing Committee on Vaccinations (STIKO) with a schedule of three priming doses in infancy plus one booster (3+1) [9]. The recommendation was extended to the entire infant population (<2 years of age) in July of 2006 [10].

PCV13 and PCV10 replaced PCV7 and were introduced in 2009, and administered based on physician’s choice [11]. In 2015, STIKO changed its recommendations for full-term infants from a 3+1 schedule with the priming series administered at 2, 3, and 4 months and a booster at 11 to 14 months, to a 2+1 vaccination schedule with a priming series administered at 2 months and 4 months plus a booster at 11 months [2, 12]. The 3+1 schedule remains in place for preterm infants [13].

PCV15 and PCV20 – next-generation PCVs with increased serotype coverage – were approved for adults aged 18 years and older by the European Commission in October of 2021 and February of 2022, respectively [14, 15]. Since September 2023, PCV20 is recommended in Germany for all individuals aged 60 years and older and for all individuals aged 18-59 years with underlying diseases. A cost-effectiveness analysis (CEA) in the German adult population concluded that a single dose of PCV20 for adults aged ≥60 years and adults aged 18-59 years with moderate– and high-risk conditions would prevent pneumococcal disease cases, save lives, and would be cost-saving compared to the pneumococcal polysaccharide vaccine (PPSV23) alone, PCV13 followed by PPSV23, or PCV15 followed by PPSV23[16].

Prior to the licensure of PCV15 in October of 2022, PCV13 was considered as the standard of care. With the inclusion of PCV15 in STIKO recommendation for infants in Germany, the current clinical practice includes a market basket of PCV13 and PCV15. PCV20 covers all PCV15 serotypes (1, 3, 4, 5, 6A, 6B, 7F, 9V, 14, 18C, 19A, 19F, 22F, 23F and 33F) and five additional serotypes (8, 10A, 11A, 12F, and 15B). On January 25th, 2024, the Committee for Medicinal Products for Human Use adopted a positive opinion for the higher-valent option – PCV20.[17] The Licensure is expected soon. The purpose of this CEA was to examine the health benefits and costs of implementing a PCV20 vaccination program under a 3+1 schedule in Germany’s pediatric population compared with PCV13 and PCV15, both administered in a 2+1 schedule.

## Methods

### Conceptual framework and model structure

The CEA was structured in Microsoft Excel^®^ (Redmond, WA, US) using a decision-analytic Markov (state-transition) cohort model. The Markov model estimated pneumococcal disease-related events in both unvaccinated and vaccinated individuals (**Figure 1**). The model captured an individual’s possible transition to several clinical events, including IPD (developing into either meningitis or sepsis/bacteremia), all-cause pneumonia (non-hospitalized or hospitalized), all-cause OM, no pneumococcal disease state, and death. Death captured both general mortality and case fatality, could occur in any disease state and non-disease state. The transition occurred on an annual cycle and were age– and vaccination-specific. The non-mutually exclusive nature of pneumococcal disease was reflected through each 12-month interval during which persons could transition to one or more disease states or remain in a non-disease state. In the case of more than one pneumococcal disease, costs and quality-adjusted life year (QALY) decrements associated with all events were considered. At the beginning of each annual cycle, a new cohort of children (i.e. incoming birth cohort) entered the model and was eligible for vaccination.

**Figure 1:**
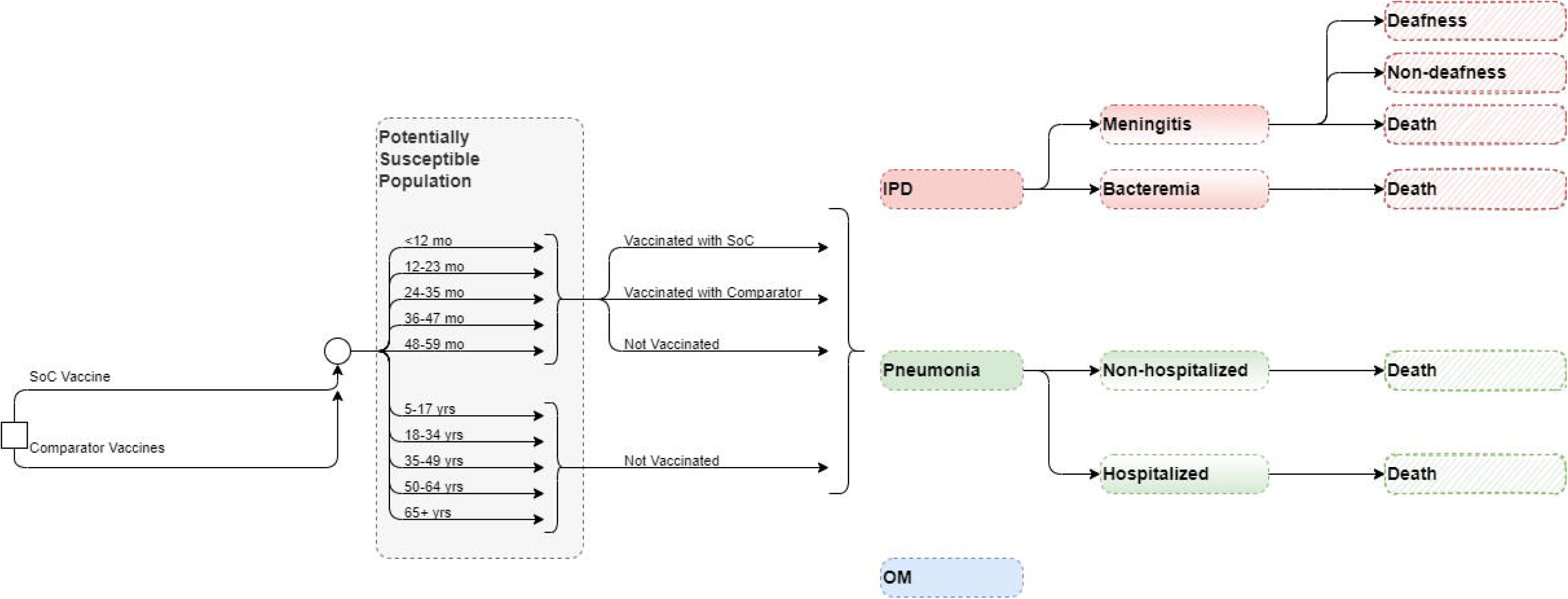
Model structure. Abbreviations: IPD, invasive pneumococcal disease; mo, months; OM, otitis media; SoC, standard of care; yrs, years

The full health benefit of vaccination was applied to the entire German population, of which the vaccinated cohort experienced the direct effects of vaccination immediately while the rest of the unvaccinated population gradually received indirect effects over the model time horizon.

### Target population and subgroups

The target population was composed of infants aged <2 years (i.e. a vaccination cohort), while the model assumed that the groups aged 2 to 4 years, 5 to 17 years, and 18 to 49 years were not vaccinated with the higher-valent PCVs. In clinical practice, a proportion of the group ≥60 years is vaccinated under the adult immunization program [18, 19], hence, within this pediatric model, that proportion of adults was excluded from receiving indirect effects, while the remainder of the population ≥60 years remained unvaccinated and benefitted from indirect effects of pediatric vaccination. The base-case population was based on German population size [20], and the sizes of incoming birth cohorts were calculated by assuming a birth rate of 1.4 children for each female [20].

Differences in event probabilities, utilities, costs, mortality, vaccine effects, and dosing schedule were reflected by stratification into several age groups (for more details, see appendix).

### Intervention and comparator strategies

STIKO currently recommends PCV10, PCV13, and PCV15 for infants and children in Germany [21]. However, PCV13 was shown to avoid more cases than PCV10 [Kuhlmann 2017] and accounts for the majority of vaccination rate, at more than 90%, in children, remaining the most used PCV in the past decade in Germany[12, 22]. Therefore, PCV10, although included in STIKO recommendation, was not considered among comparators in this analysis. The analysis evaluated the clinical and economic outcomes of PCV20 in a 3+1 schedule as a potential vaccination strategy compared with PCV13 (i.e., SoC) and PCV15, both in a 2+1 schedule.

### Perspective, time horizon, cycle length, and discount rate

The base-case analysis was conducted from a German societal perspective using a 3% annual discount rate for both costs and benefits, according to the recommendations of the Institute for Quality and Efficiency in Health Care and STIKO [23, 24]. The model used an annual cycle length over a 10-year time horizon to capture relevant costs and outcomes. The 10-year time horizon sufficiently captures the health benefits of the PCV vaccination program, based on the observation of the accrual and stabilization of indirect effects over a five-to 10-year period following the introduction of PCV7 and PCV13 [25, 26]. The life years and QALY loss is accumulated over the time loss between death occurrence and life expectancy (with the QALYs being age dependent and discounted from the year the death occurs). Lifetime long-term costs related to a clinical event such as sequalae following meningitis were incorporated in the model as a one-time discounted cost in the cycle in which the event happens.

### Inputs

#### Population and epidemiology data

Population data were obtained from the German Federal Statistical Office to determine the size of the stratified population age groups [27] (**Table S1**) and to inform the assumption of a birth rate of 1.4 per female to estimate the number of infants in the incoming birth cohort each year [20] (**Table S2**).

Age-specific disease incidence rates per 100,000 were informed by German-specific published literature [28–31]. If the age groups did not align with those in the model, the incidence rates were adjusted using data from the literature and population size from the German Federal Statistical Office [27] (**Table 1**Error! Reference source not found.). IPD cases were assumed to develop into either meningitis or sepsis/bacteremia.

**Table 1:** Epidemiology inputs.

|  | Disease incidence per 100,000 |  |  |  | Breakdown of IPD cases<br>[20, 72] |  | Case fatality rate |  |  |  |
| --- | --- | --- | --- | --- | --- | --- | --- | --- | --- | --- |
|  | IPD [28, 29] | Hospitalized pneumonia [29, 33] | Non-hospitalized pneumonia [29, 33] | Otitis media [31] | Meningitis | Sepsis/bacteraemia | Meningitis [29, 73] | Sepsis/bacteraemia [29, 73] | Hospitalized pneumonia [29, 73] | Non-hospitalized pneumonia [74] |
| <12 months | 15.8 | 1,493 | 2,447 | 14,749 | 33.40% | 66.60% | 6.80% | 1.00% | 0.15% | 0.00% |
| 12–23 months | 15.8 | 814 | 2,447 | 14,749 | 33.40% | 66.60% | 10.70% | 2.20% | 0.09% | 0.00% |
| 24–47 months | 1.4 | 814 | 8,492 | 17,939 | 33.40% | 66.60% | 6.90% | 2.30% | 0.09% | 0.00% |
| 48–59 months | 1.4 | 814 | 8,492 | 3,794 | 33.40% | 66.60% | 6.90% | 2.30% | 0.09% | 0.00% |
| 5–17 years | 1.0 | 132 | 2,426 | - | 33.40% | 66.60% | 4.81% | 7.23% | 0.90% | 0.00% |
| 18–49 years | 1.7 | 111 | 401 | - | 9.10% | 90.90% | 8.74% | 8.74% | 4.42% | 0.00% |
| 50–64 years | 10.6 | 538 | 691 | - | 4.60% | 95.40% | 12.95% | 12.95% | 11.94% | 0.40% |
| ≥65 years | 22.0 | 2,550 | 1,022 | - | 1.80% | 98.20% | 19.65% | 19.65% | 18.67% | 0.40% |

Abbreviation: IPD, invasive pneumococcal disease

Mortality in the analysis was considered as a combination of general mortality [32] and case fatality, which were applied to meningitis, sepsis/bacteremia, and all-cause hospitalized and non-hospitalized pneumonia (**Table 1**), while no mortality was assumed for OM [29, 33].

The model considered sequela following meningitis (i.e., deafness and non-deafness) in the base case to capture the disease burden related to IPD as sequela following meningitis are quite common among patients with IPD [33]. The data for the proportion of patients developing complications with IPD were sourced from Kuhlmann et al., 2017 [33] (**Table S3)**. Sequela following other pneumococcal diseases such as pneumonia and OM were not considered in this analysis.

IPD serotype-specific distribution by each PCV stratified by age groups was obtained from van der Linden and Itzek, 2022 [34, 35] **(Table 2**). Serotype distribution was calculated from the number of reported cases due to each individual PCV serotype specified by age groups. The serotype coverage for PCV7 serotypes (4, 6B, 9V, 14, 18C, 19F, 23F) was incorporated as one input, while the coverage for each additional serotype included in higher valent vaccines was input separately for each age group. The percentage of non-vaccine covered serotypes equaled 100% minus all vaccine-type serotype coverage. The analysis did not consider cross-reactive serotypes. Non-invasive serotype distributions (i.e. for pneumonia and OM) were assumed to be the same as IPD serotype distribution.

**Table 2:** Current serotype distribution by age. [34, 35]

| Age | PCV-7 serotypes | Serotypes additional to PCV-7 |  |  |  |  |  |  |  |  |  |  |  |  | Non-vaccine covered |
| --- | --- | --- | --- | --- | --- | --- | --- | --- | --- | --- | --- | --- | --- | --- | --- |
|  |  | PCV-10 |  |  | PCV-13 |  |  | PCV-15 |  | PCV-20 |  |  |  |  |  |
|  |  | 1 | 5 | 7F | 3 | 6A | 19A | 22F | 33F | 8 | 10A | 11A | 12F | 15B |  |
| <23 months | 5.9% | 0.0% | 0.0% | 0.0% | 4.0% | 1.0% | 2.0% | 2.0% | 3.0% | 3.0% | 15.8% | 0.0% | 5.0% | 5.9% | 52% |
| 24–59 months | 5.1% | 0.0% | 0.0% | 0.0% | 5.1% | 0.0% | 10.3% | 7.7% | 0.0% | 0.0% | 10.3% | 5.1% | 5.1% | 12.8% | 38% |
| 5–17 years | 5.4% | 0.0% | 0.0% | 0.0% | 10.8% | 0.0% | 13.5% | 10.8% | 0.0% | 10.8% | 2.7% | 8.1% | 5.4% | 2.7% | 30% |
| 18–49 years | 10.4% | 0.5% | 0.3% | 0.8% | 12.0% | 0.0% | 3.7% | 5.6% | 0.8% | 20.9% | 1.6% | 2.4% | 8.3% | 1.3% | 31% |
| 50–64 years | 5.0% | 0.0% | 0.0% | 0.3% | 20.8% | 0.2% | 4.0% | 7.3% | 1.2% | 16.4% | 2.4% | 2.7% | 5.1% | 1.5% | 33% |
| ≥65 years | 3.7% | 0.1% | 0.0% | 0.3% | 23.1% | 0.4% | 3.3% | 7.9% | 1.5% | 7.8% | 3.8% | 3.8% | 3.5% | 1.8% | 39% |

Abbreviation: PCV, pneumococcal conjugate vaccine

#### Vaccine effectiveness and efficacy

The direct effect of PCVs against IPD was estimated using data from Savulescu et al., 2022 [36], which aligned with the approach in Lytle et al., 2023 [37]. For a complete vaccine schedule, the model assumed the effectiveness of higher-valent vaccines against vaccine-type IPD would be equivalent to the adjusted PCV13 effectiveness against PCV13-type IPD as reported in Savulescu et al., 2022, of which 78.2% (95% confidence interval CI: 56.0, 89.0) was applied for vaccines in a 2+1 schedule while 89.7% (95% CI: 82.0, 94.0) was used for PCV20 in 3+1 schedule (**Table 3)**. To estimate the direct effects of the higher-valent PCVs against all-cause pneumonia (non-hospitalized and hospitalized) and OM, the model adopted an approach commonly used in CEAs [38–42] in which the effectiveness of higher-valent PCVs against non-invasive disease is assumed to be the same as the reported trial-based efficacy data of PCV7, which was then adjusted based on study design, period, and country-specific factors. These results demonstrated an efficacy of 25.5% (95% CI: 4.4, 34.0) [43], 6.0% (95% CI: –1.5, 11.0) [44], and 7.8% (95% CI: 5.2, 10.5) [45] against radiographically confirmed non-invasive hospitalized pneumonia, non-hospitalized pneumonia, and OM, respectively (**Table 3**).

**Table 3:**
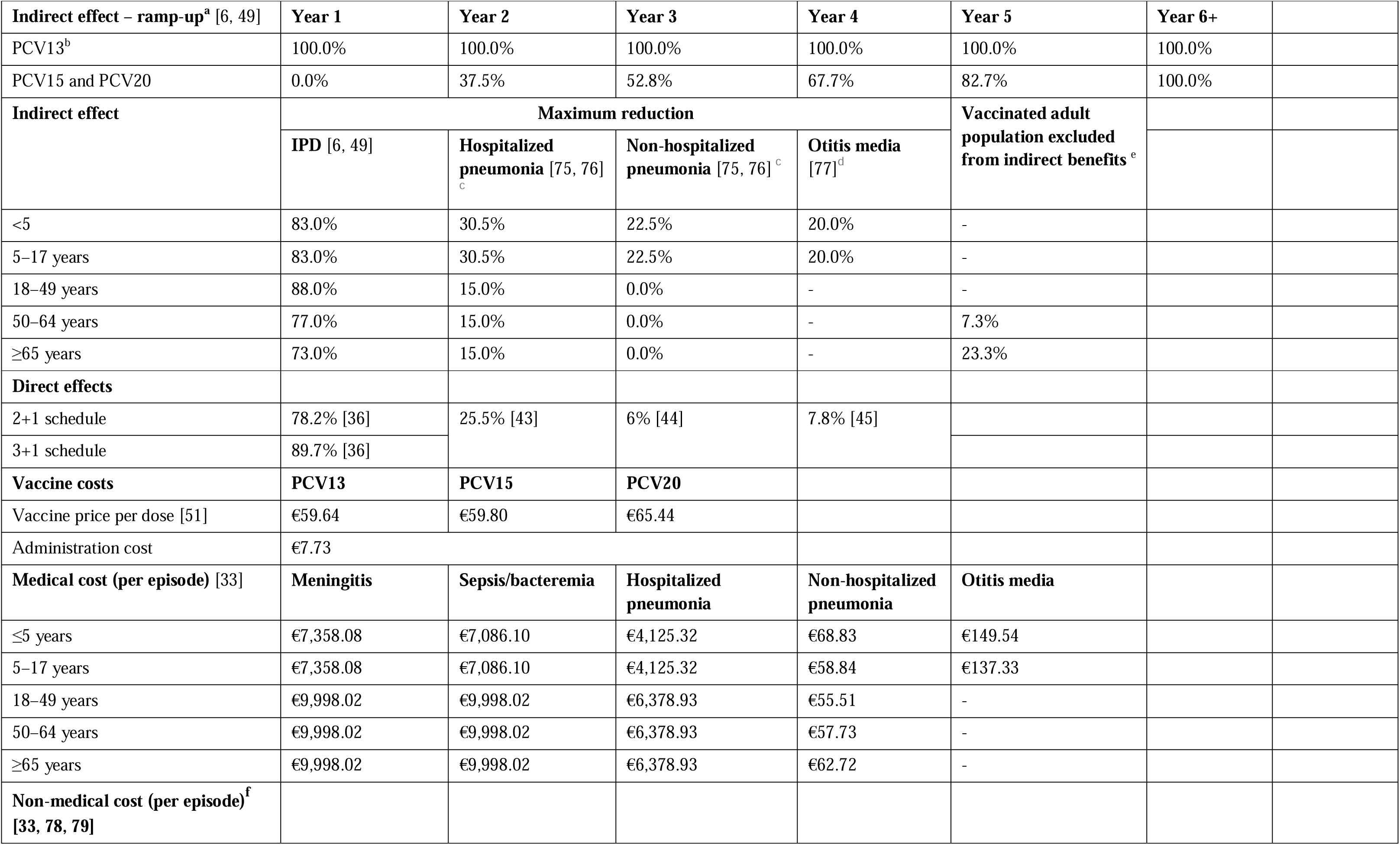
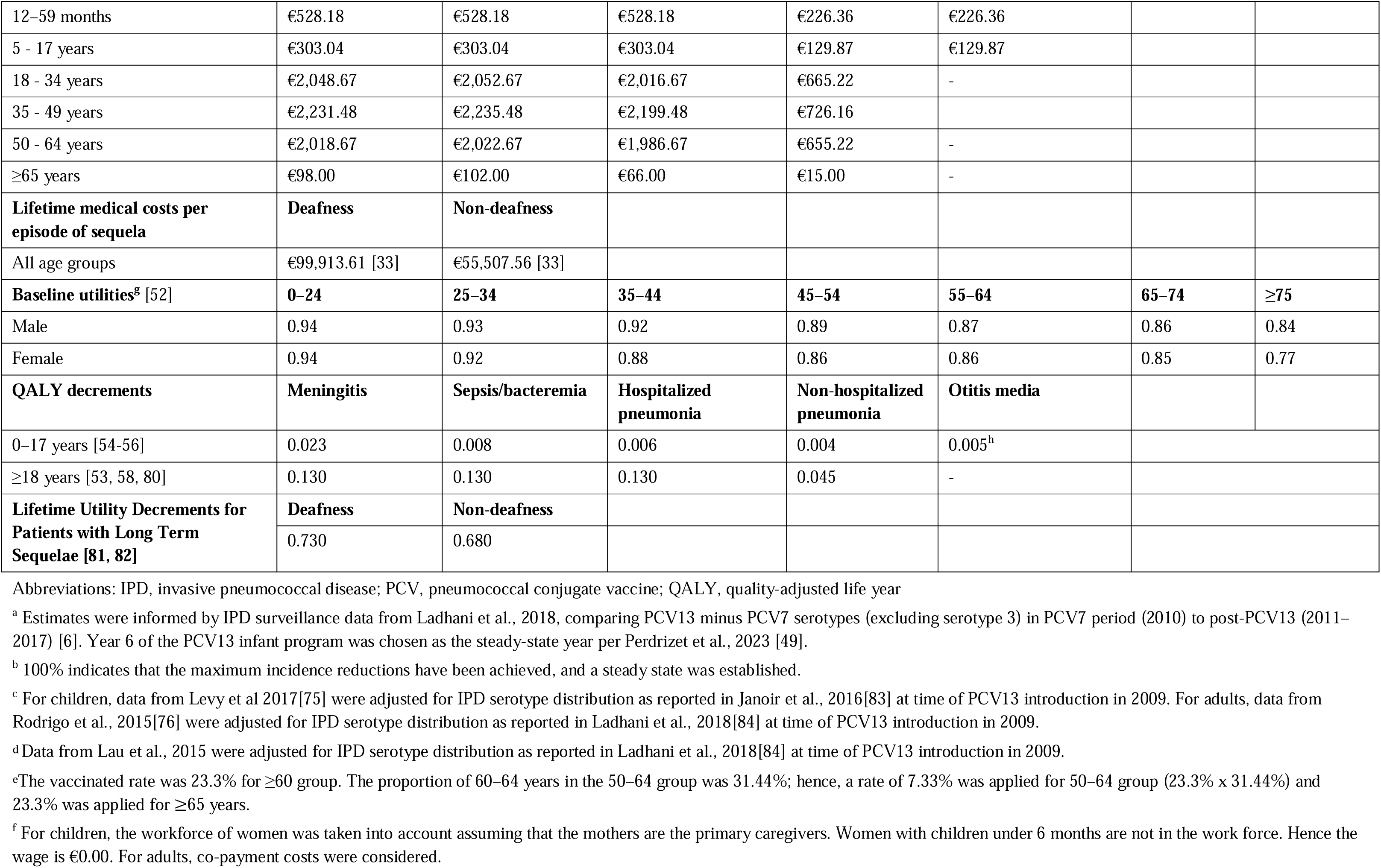
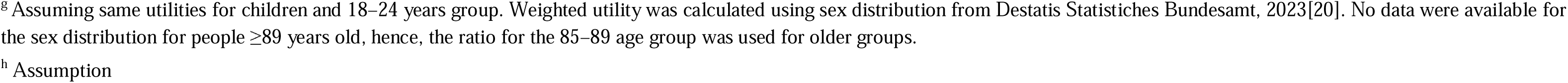
Vaccine effectiveness, cost, and utility parameters.

In addition, a <12-month effect modifier was used to account for potential reduced effectiveness in the first year of life during which children have only received the priming series of the full vaccination schedule. The <12-month modifier was set at two-thirds (∼67%) of the full effect for vaccination with a 2+1 schedule (PCV13 and PCV15) and at 75.6% for PCV20 3+1 based on the Advisory Committee on Immunization Practices’s assumption by Stoecker 2021 [46]. Vaccine coverage was set at 89.90% for the priming series and at 76.80% for the booster dose [47]. The booster dose is given at month 14 for PCV20 and at month 11 for both PCV13 and PCV15. However, real-world data from 2013 to 2018 for German clinical practice showed that the highest proportion of boosters were given in month 14 (∼20%), so the model assumed all booster doses are administered at 14 months [47].

Evidence has shown that direct effects of PCVs remain stable for a few years after the final dose. For example, the efficacy of PCV13 was steady for up to four years in infants after vaccination was completed [36] and for more than five years in people aged ≥65 years after a single dose [48]. Therefore, in the base case, a full direct effect for the first five years after the booster dose was assumed for all vaccines, followed by an annual waning of 10% from year 6 through year 10.

The analysis considered indirect effects in unvaccinated individuals since they are an important benefit from pediatric PCV national immunization programs. The indirect effect against serotypes covered represents the maximum protection the unvaccinated population could receive from a vaccine regimen. This was modelled as a percent reduction in the expected age-specific disease incidence. Indirect effect was not realized immediately and was only applied to newly covered serotypes in PCV15 and PCV20 as the indirect effect for PCV13 serotypes was assumed to have already reached a steady state. These benefits accrued gradually until a new steady state was reached for additional serotypes. Indirect effect for PCV15 and PCV20 was assumed to have no added effects on PCV13 steady-state serotypes. The model assumed that incidence trends for all newly covered serotypes would decrease consistently across ages. For IPD, non-hospitalized and hospitalized pneumonia, indirect effect was assumed for all age groups, while for OM, indirect effect was assumed only for the <5 years age group.

To estimate indirect effects, the model incorporated the reduction in incidence of the newly covered serotypes and the accrual of the indirect effects of higher-valent PCVs (see Appendix A). The accrual of the indirect effects of higher-valent PCVs was informed by IPD surveillance data from Ladhani et al., 2018 [6] that compared PCV13 minus PCV7 serotypes (excluding serotype 3) and from year 6 onward of the PCV13 infant program when the steady state was reached (Perdrizet et al., 2023[49]). Input data for the maximum reduction in disease incidence estimated separately for relevant age groups from several impact studies are summarized in **Table 3**. As children aged <2 years will benefit from both direct effect and indirect effect, the model followed an impact approach where data from PCV13 impact studies for subjects aged 5-17 years (i.e. pure indirect effects from PCV13) were used for <2-year-old cohort to avoid double counting.

#### Resource use and costs

The model considered the costs for vaccine doses, administration, and medical resource use, as well as lifetime medical costs per episode of sequela. All costs were in Euros (€) and obtained from German published sources and literature, then inflated to 2022 prices using the healthcare component of the consumer price index [50] where relevant.

Vaccine costs for PCV13, PCV20, and PCV15 were derived from retail pharmacy price per dose [51] and the administration cost were from Kuhlmann et al., 2017[33], inflated to 2022 Euro. Additional vaccine cost and administration cost were applied to the extra dose under 3+1 schedule for PCV20. Medical costs per episode related to each disease state sourced from Kuhlman et al., 2017 were included for all relevant age groups in the model [33]. Lifetime medical costs per episode of sequela were assumed to be the same across all age groups for deafness and non-deafness [33]. Societal costs considered productivity loss per episode of meningitis, sepsis/bacteremia, inpatient and outpatient pneumonia and OM, as well copayment for adult patients. The summary of cost inputs is listed in **Table 3**.

#### Utility

The model used baseline utility for the general population [52] minus disutilities related to disease states and acute events to assess quality of life related to each vaccination strategy. QALY decrements were sourced from the literature for IPD (meningitis and sepsis/bacteremia), hospitalized and non-hospitalized pneumonia, and OM (**Table 3**) [42, 53–58].

### Assessment of uncertainty

Uncertainty around the analyses was evaluated using deterministic sensitivity analyses (DSA), probabilistic sensitivity analyses (PSA), and scenario analyses. The DSA used the 95% CI of each parameter where available, or a 20% proxy variation to estimate upper and lower bound of inputs and applied them as new inputs into the model. DSA assessed uncertainty around the following variables: disease incidence, breakdown of IPD cases, CFR, serotype distribution by age, vaccine effectiveness and utilities.

To calculate the PSA, all parameters subject to any degree of uncertainty were assessed using probability distributions following Briggs et al., (2006), from which a random value could be drawn [59]. The variations of relevant parameters, such as incidence within a certain disease state across different age groups or CFR stratified by age, were controlled to vary in the same direction to ensure consistency between those variables that are not completely independent of one another. The incremental results for costs, QALYs, and incremental cost-effectiveness ratios (ICER) were recorded for each simulation of a total of 1,000 simulations to examine the stability of the model findings.

Several scenarios were conducted around discount rates for costs and outcomes (i.e., no discount for both costs and outcomes, 0% and 1.5% discount rates applied for outcomes while 3% is applied for costs) to examine the structural uncertainty of the model. Different assumptions in vaccine indirect effects were tested, such as uniformly reducing indirect effects by half for all diseases and age groups, extending the ramp-up duration to two years from the last dose of vaccine to start realizing the indirect effects and increasing the vaccine uptake in adults. Since waning was based on assumptions, a different assumption was tested by reducing duration of full protection to three years (i.e., waning completed by year 8). Three scenarios were considered with serotype unmasking: the proportion of newly covered serotype was assumed to reduce annually by 5% (1) and 10% (2) compared to baseline for five years, after which they were assumed to remain in steady state and the proportion of newly covered serotypes was assumed to reduce by the same percentages that PCV13minus PCV7 serotype disease decreased following PCV13 implementation in Germany (3) until a steady state was reached. Payer perspective and an assumed 90% vaccine uptake were also tested. Finally, the model considered a scenario where disutility related the administration of vaccines was applied for all PCVs.

## Results

### Base-case results

Over the 10-year time horizon, compared to both PCV13 2+1 and PCV15 2+1, PCV20 3+1 provided substantially greater health benefits and broader protection **(Table 4)**. Compared to PCV13, PCV20 is estimated to result in greater health benefits due to a greater number of cases averted and total QALYs gained **(Table 4)**. Compared with PCV13, PCV20 averted an additional 15,301 cases of IPD; 460,197 and 472,365 cases of hospitalized and non-hospitalized pneumonia respectively; 531,634 cases of OM; and 59,265 deaths due to disease across all ages. Consequently, PCV20 was estimated with a higher QALY gain of 904,854. In comparison with PCV15, the number of additional cases averted by PCV20 were 11,334, 335,937, 369,012, 441,643 in IPD, hospitalized and non-hospitalized pneumonia and OM in turn. Additionally, an estimation of a reduction of 41,596 deaths due to disease was estimated for PCV20 versus PCV15 and PCV20 was associated with an incremental QALY of 646,235.

**Table 4:** Cost-effectiveness results and incremental difference of PCV20 vs PCV13 and PCV20 vs PCV15.

| Outcome | PCV13 2+1 | PCV15 2+1 | PCV20 3+1 | PCV20 vs. PCV13 | PCV20 vs. PCV15 |
| --- | --- | --- | --- | --- | --- |
| Total pneumococcal cases | 22,306,443 | 21,984,871 | 20,826,946 | -1,479,497 | -1,157,925 |
| Cases of IPD | 77,013 | 73,046 | 61,712 | -15,301 | -11,334 |
| Cases of hospitalized pneumonia | 7,512,228 | 7,387,967 | 7,052,031 | -460,197 | -335,937 |
| Cases of non-hospitalized pneumonia | 9,498,563 | 9,395,209 | 9,026,198 | -472,365 | -369,012 |
| Cases of otitis media | 5,218,640 | 5,128,648 | 4,687,006 | -531,634 | -441,643 |
| Number of deaths due to disease | 1,112,503 | 1,094,834 | 1,053,238 | -59,265 | -41,596 |
| Total costs | €49,750,634,550 | €48,985,371,445 | €47,357,370,939 | -€2,393,263,611 | -€1,628,000,506 |
| Cost of vaccination (doses and administration) | €1,100,550,473 | €1,103,164,936 | €1,625,912,755 | €525,362,283 | €522,747,819 |
| Medical costs | €42,742,979,399 | €42,051,612,831 | €40,194,950,206 | -€2,548,029,193 | -€1,856,662,625 |
| Costs of lifetime sequelae | €48,755,844 | €46,219,830 | €36,295,226 | -€12,460,617 | -€9,924,603 |
| Indirect cost of disease | €5,858,348,835 | €5,784,373,848 | €5,500,212,751 | -€358,136,083 | -€284,161,097 |
| Total LYs | 2,024,138,619 | 2,024,300,902 | 2,024,701,633 | 563,014 | 400,731 |
| Total QALYs | 1,741,648,852 | 1,741,907,472 | 1,742,553,707 | 904,854 | 646,235 |
| ICER per QALY | - | - | - | Dominant | Dominant |

Abbreviations: ICER, incremental cost-effectiveness ratio; IPD, invasive pneumococcal disease; LYs, life years; OM, otitis media; PCV, pneumococcal conjugate vaccine; QALYs, quality-adjusted life years; SoC, standard of care

Although PCV20 was associated with higher direct vaccine costs, it resulted in significant cost savings from lower direct costs of disease and lifetime costs of sequela compared to both comparators **(Table 4)** due to broader serotype coverage compared to PCV13 and PCV15. PCV20 was the dominant strategy in both comparisons, with a total cost saving of €2,393,263,611 versus PCV13 and of €1,628,000,506 versus PCV15.

The breakdown results by age groups were mostly consistent with the overall results where more cases avoided related to PCV20 in all included age groups for IPD, hospitalized pneumonia and OM than PCV13, especially in the oldest group of 65+ year-olds. Similarly, PCV20 is estimated to prevent more cases of non-hospitalized pneumonia than PCV13 in children less than 5 years old. The higher valent vaccines showed an increasing number of prevented non-invasive pneumonia. One could observe a shift from severe cases (i.e., hospitalized cases) in the lower valent vaccine strategies to less severe cases (i.e.; non-hospitalized pneumonia) in the higher valent vaccine strategies. Furthermore, better health outcomes from PCV20 were shown by a reduction in deaths due to disease in all age groups, with the greatest reduction observed in those 65 years old and above (50,064 deaths averted) (**Table S4**). Similar results broken down by age groups were also observed when comparing PCV20 with PCV15 (**Table S4** and **Figure 2**–**Figure 5**).

**Figure 2:**
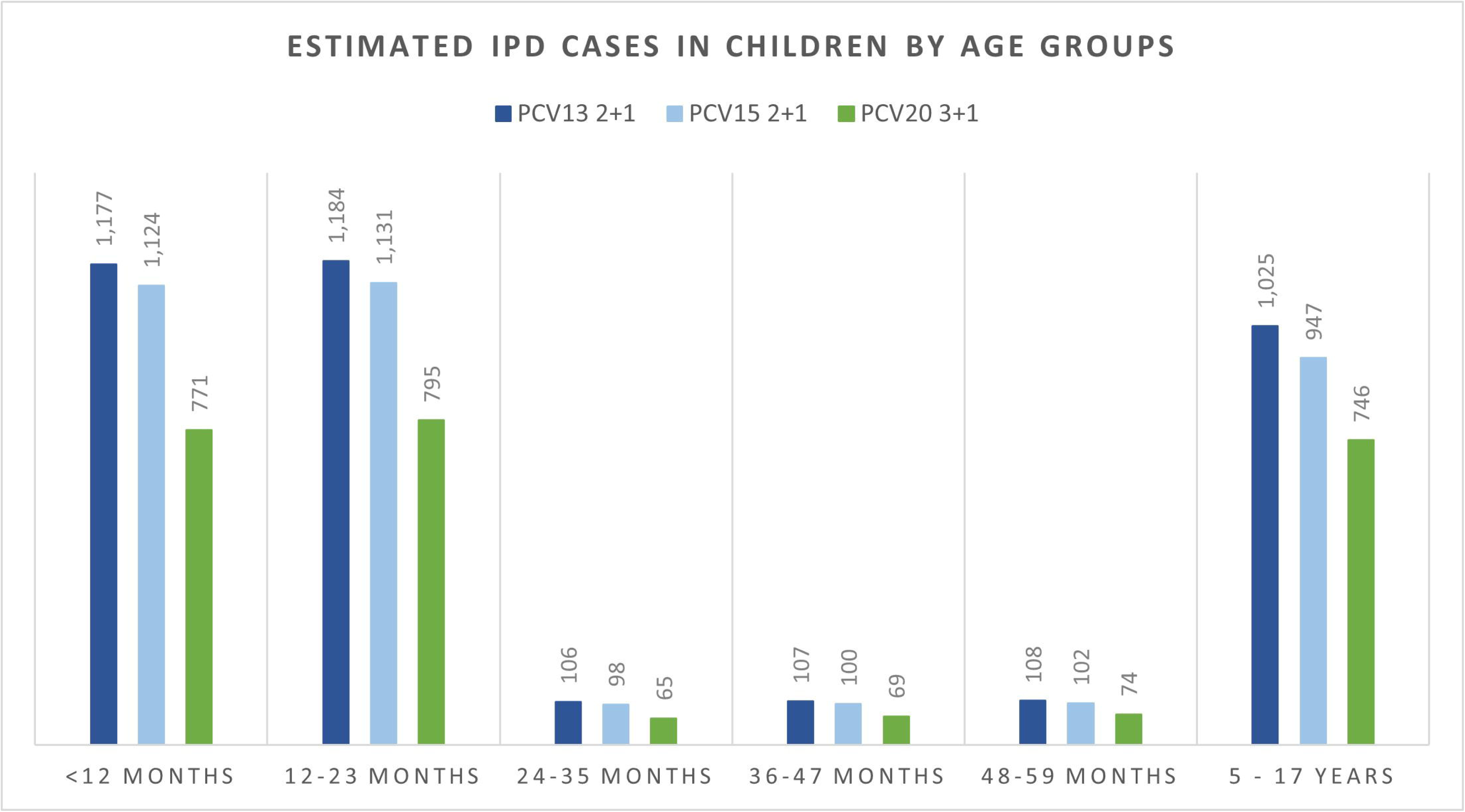
Estimated number of IPD cases stratified by age groups. Abbreviations: IPD, invasive pneumococcal disease; PCV13, 13-valent pneumococcal conjugate vaccine; PCV15, 15-valent pneumococcal conjugate vaccine; PCV20, 20-valent pneumococcal conjugate vaccine

**Figure 3:**
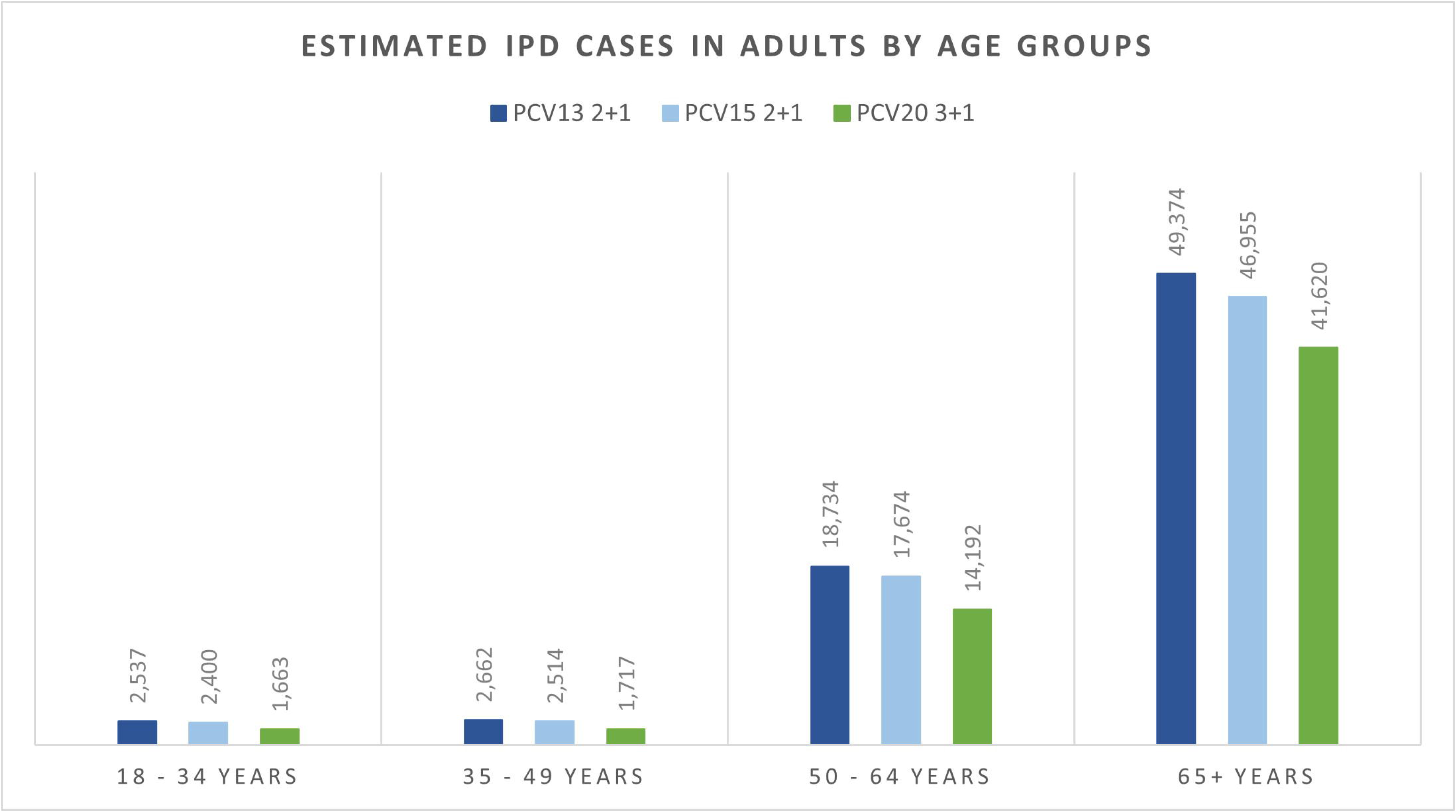
Estimated number of hospitalized pneumonia cases stratified by age groups. Abbreviations: PCV13, 13-valent pneumococcal conjugate vaccine; PCV15, 15-valent pneumococcal conjugate vaccine; PCV20, 20-valent pneumococcal conjugate vaccine

**Figure 4:**
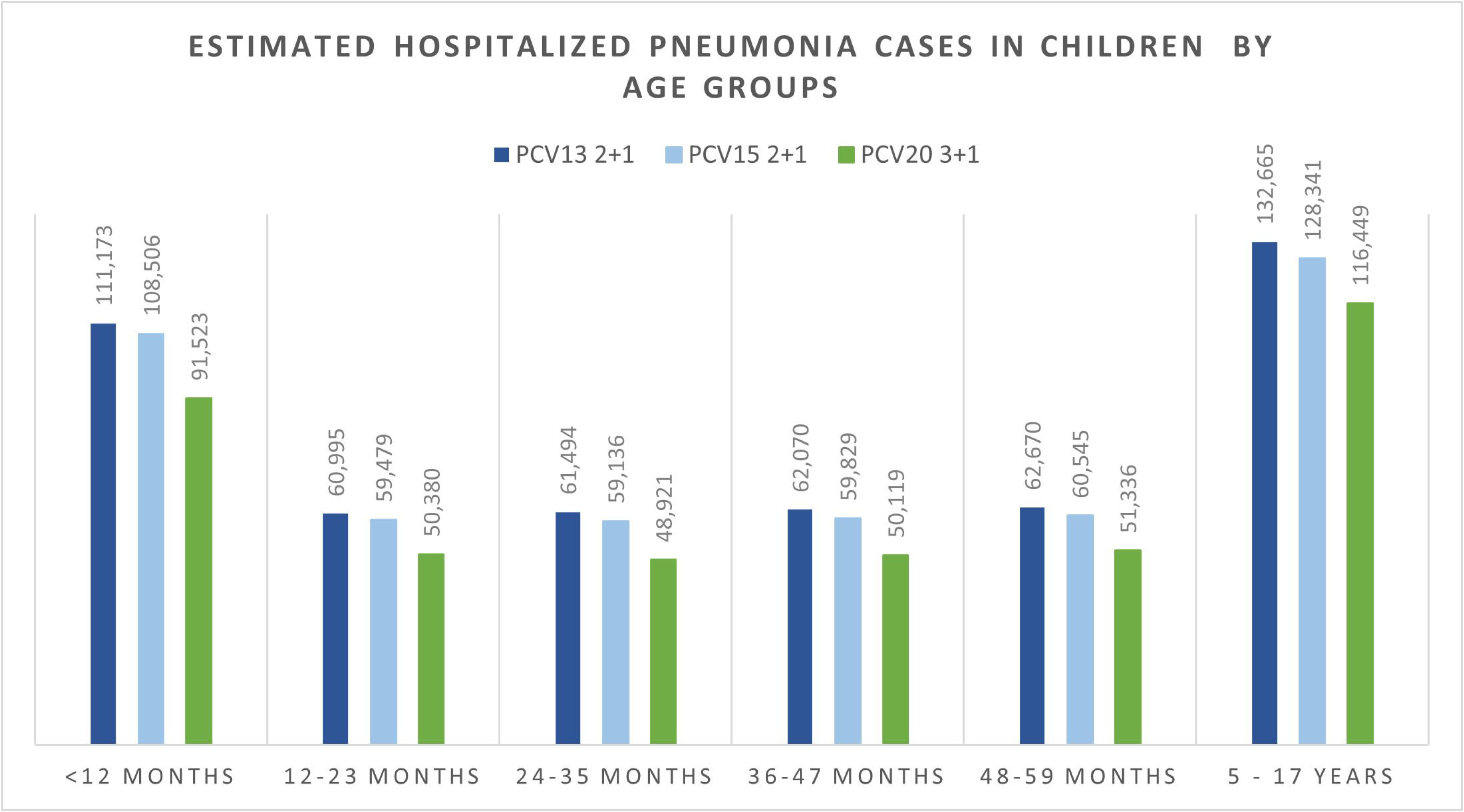
Estimated number of non-hospitalized pneumonia cases stratified by age groups. Abbreviations: PCV13, 13-valent pneumococcal conjugate vaccine; PCV15, 15-valent pneumococcal conjugate vaccine; PCV20, 20-valent pneumococcal conjugate vaccine

**Figure 5:**
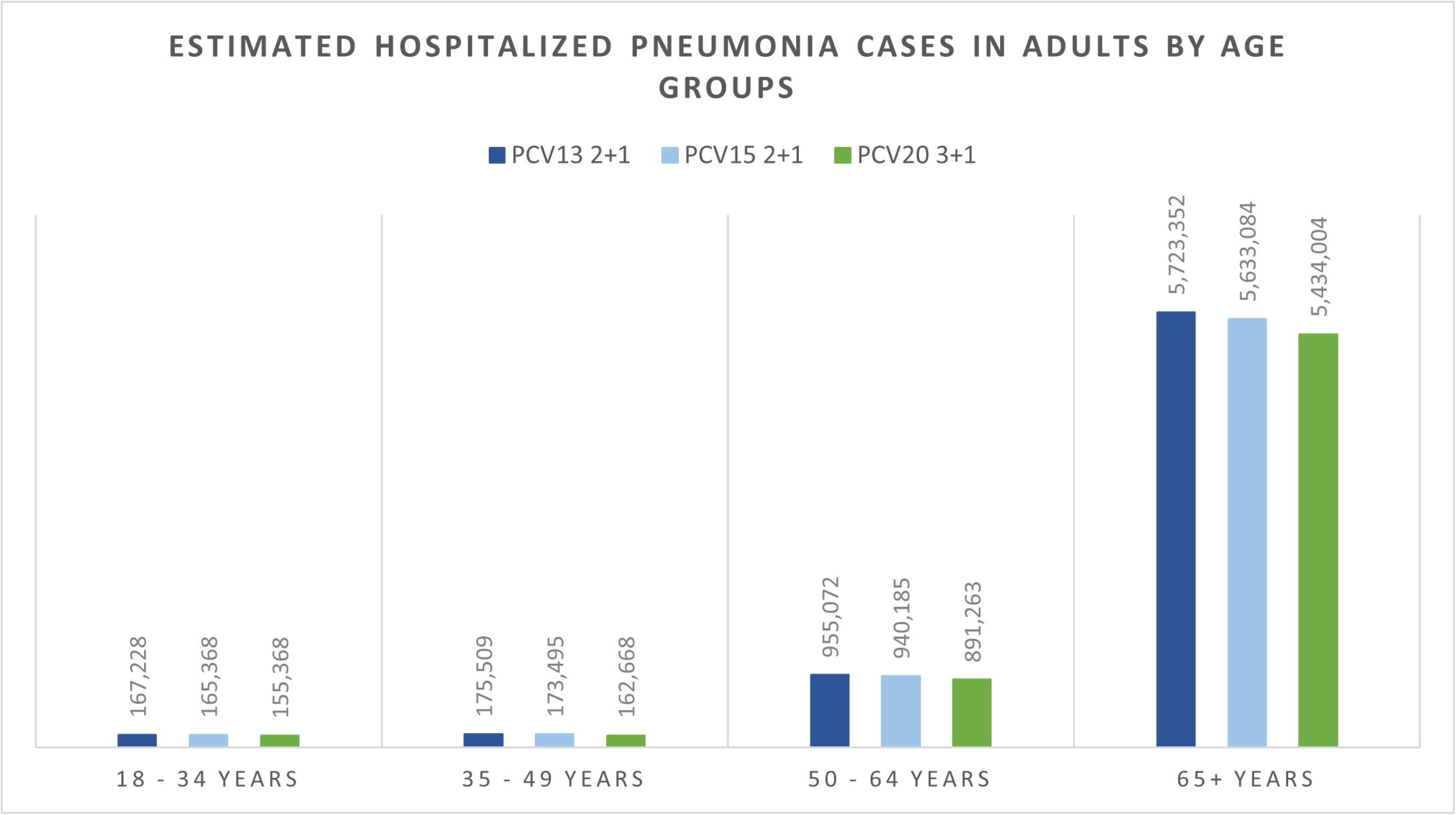
Estimated number of otitis media cases stratified by age groups. Abbreviations: PCV13, 13-valent pneumococcal conjugate vaccine; PCV15, 15-valent pneumococcal conjugate vaccine; PCV20, 20-valent pneumococcal conjugate vaccine

### Sensitivity analyses

The results from DSA, where one parameter was varied in one direction while all other inputs were held constant, are reported in **Figure 6** for costs and **Figure 7** for QALYs. When compared to PCV13, the key drivers for costs included maximum indirect effect against hospitalized pneumonia (PCV20), serotype distribution by age, incidence of hospitalized pneumonia and medical costs of per episode of hospitalized pneumonia.

**Figure 6:**
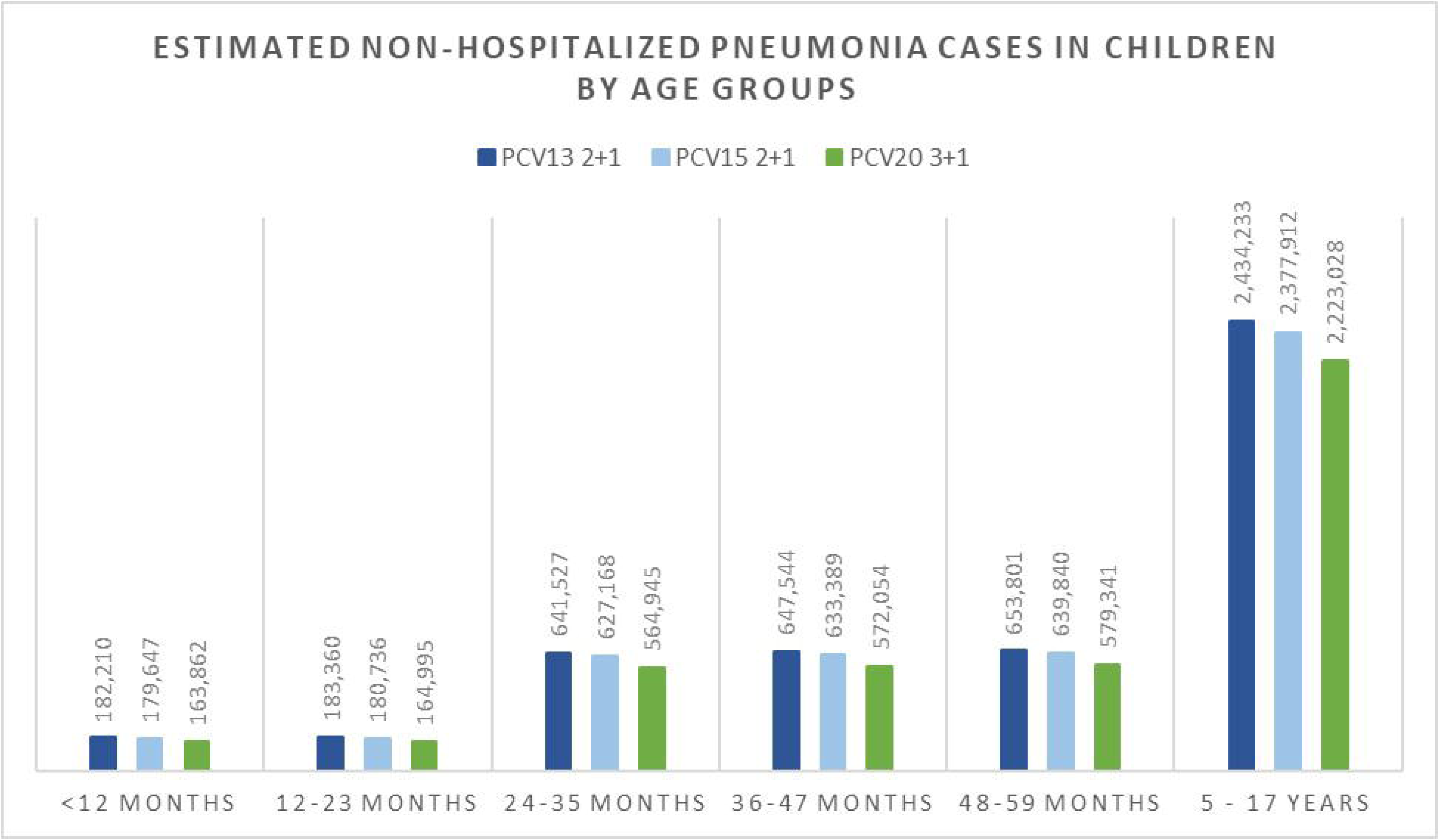
DSA results in costs: PCV20 vs PCV13. Abbreviations: DSA, deterministic sensitivity analysis; PCV13, 13-valent pneumococcal conjugate vaccine; PCV20, 20-valent pneumococcal conjugate vaccine

**Figure 7:**
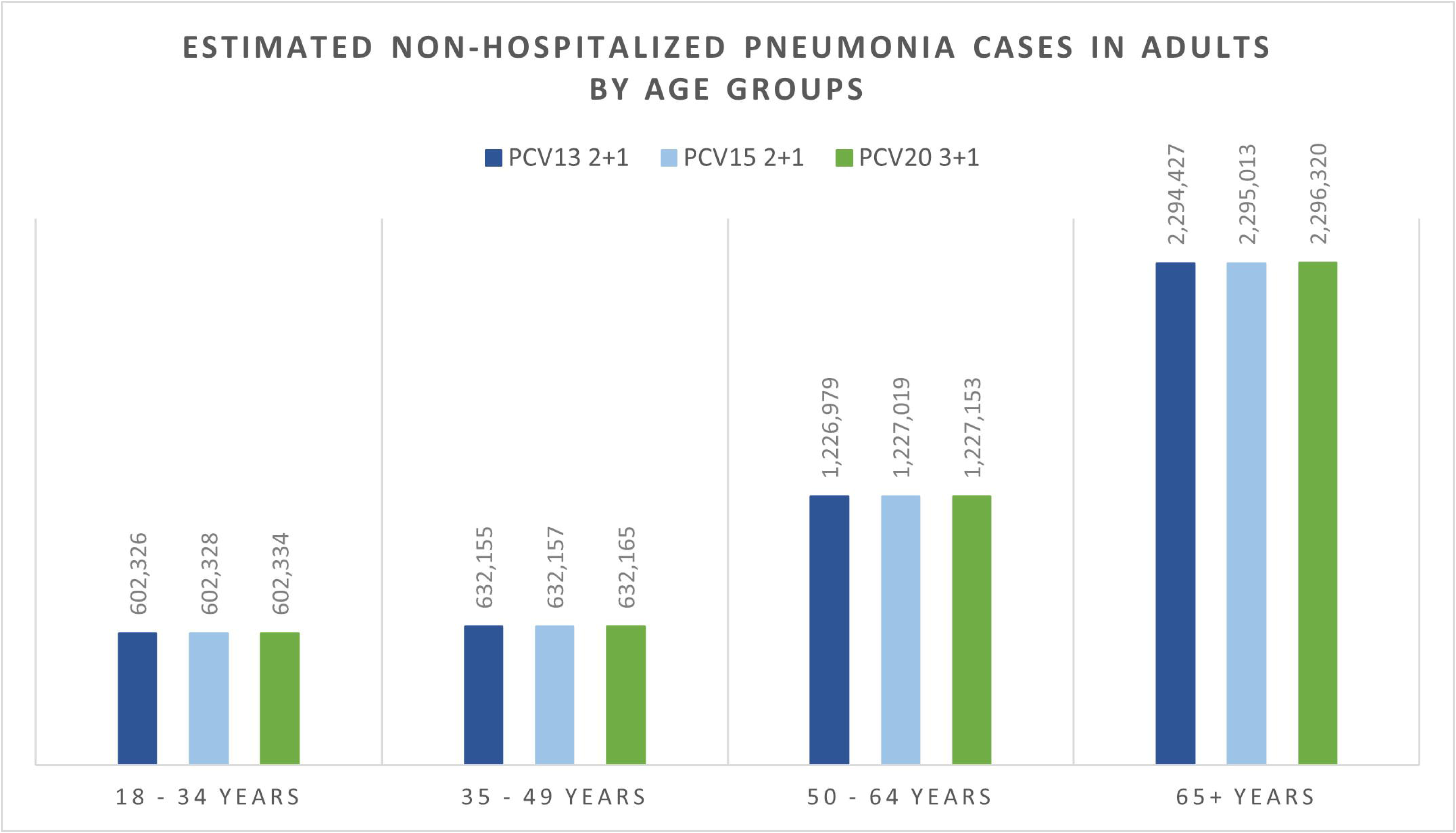
DSA results in QALYs: PCV20 vs PCV13. Abbreviations: DSA, deterministic sensitivity analysis; PCV13, 13-valent pneumococcal conjugate vaccine; PCV20, 20-valent pneumococcal conjugate vaccine; QALY, quality-adjusted life years

The DSA of PCV20 versus PCV13 also illustrated that the top five most impactful parameters on QALYs were maximum indirect effects on hospitalized pneumonia (PCV20) and serotype distribution by age, baseline utilities, followed by hospitalized pneumonia incidence and CFR for hospitalized pneumonia. When comparing PCV20 and PCV15, the results were largely similar (**Figure 8** and **Figure 9**).

**Figure 8:**
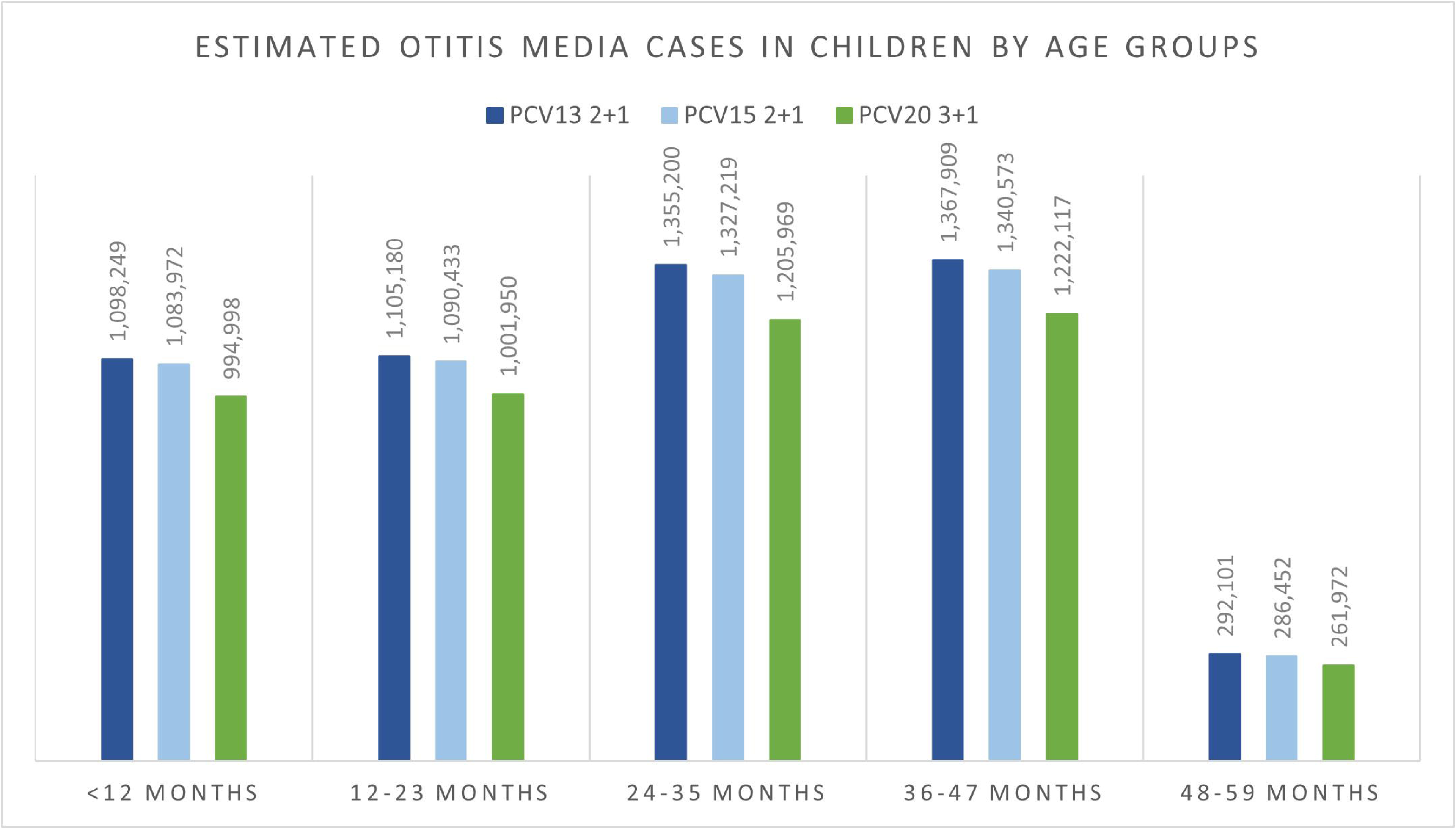
DSA results in costs: PCV20 vs PCV15. Abbreviations: DSA, deterministic sensitivity analysis; PCV15, 15-valent pneumococcal conjugate vaccine; PCV20, 20-valent pneumococcal conjugate vaccine

**Figure 9:**
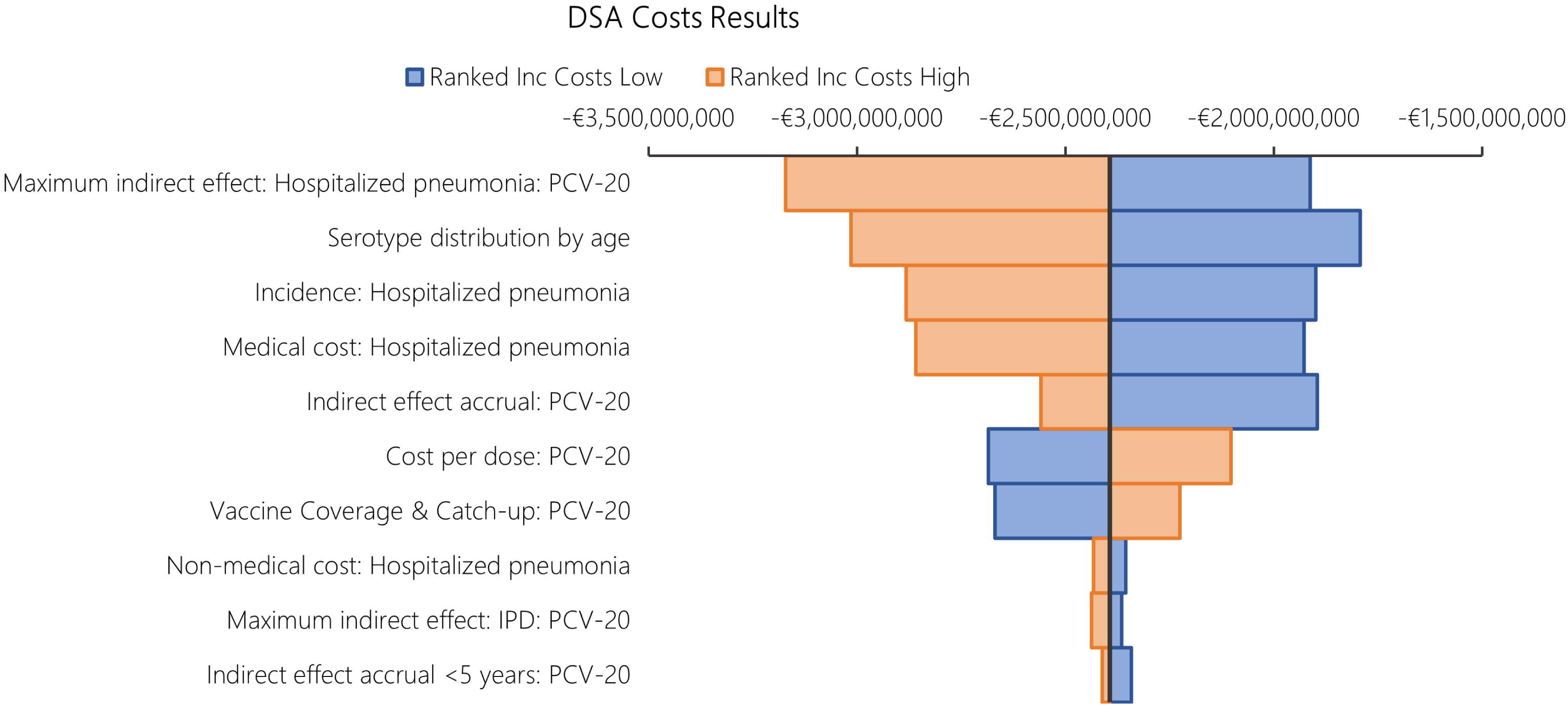
DSA results in QALYs: PCV20 vs PCV15. Abbreviations: DSA, deterministic sensitivity analysis; PCV15, 15-valent pneumococcal conjugate vaccine; PCV20, 20-valent pneumococcal conjugate vaccine; QALY, quality-adjusted life years

Probabilistic results from the PSA based on 1,000 iterations aligned with the base case results, confirming robust findings with PCV20 being dominant in all simulations. Compared with PCV13, PCV20 was the dominant strategy in all iterations, while PCV20 dominated PCV15 in 98.40% of the total 1000 iterations **(Table 5).** The cost-effectiveness plane plots are reported in **Figure 10** and **Figure 11**.

**Figure 10:**
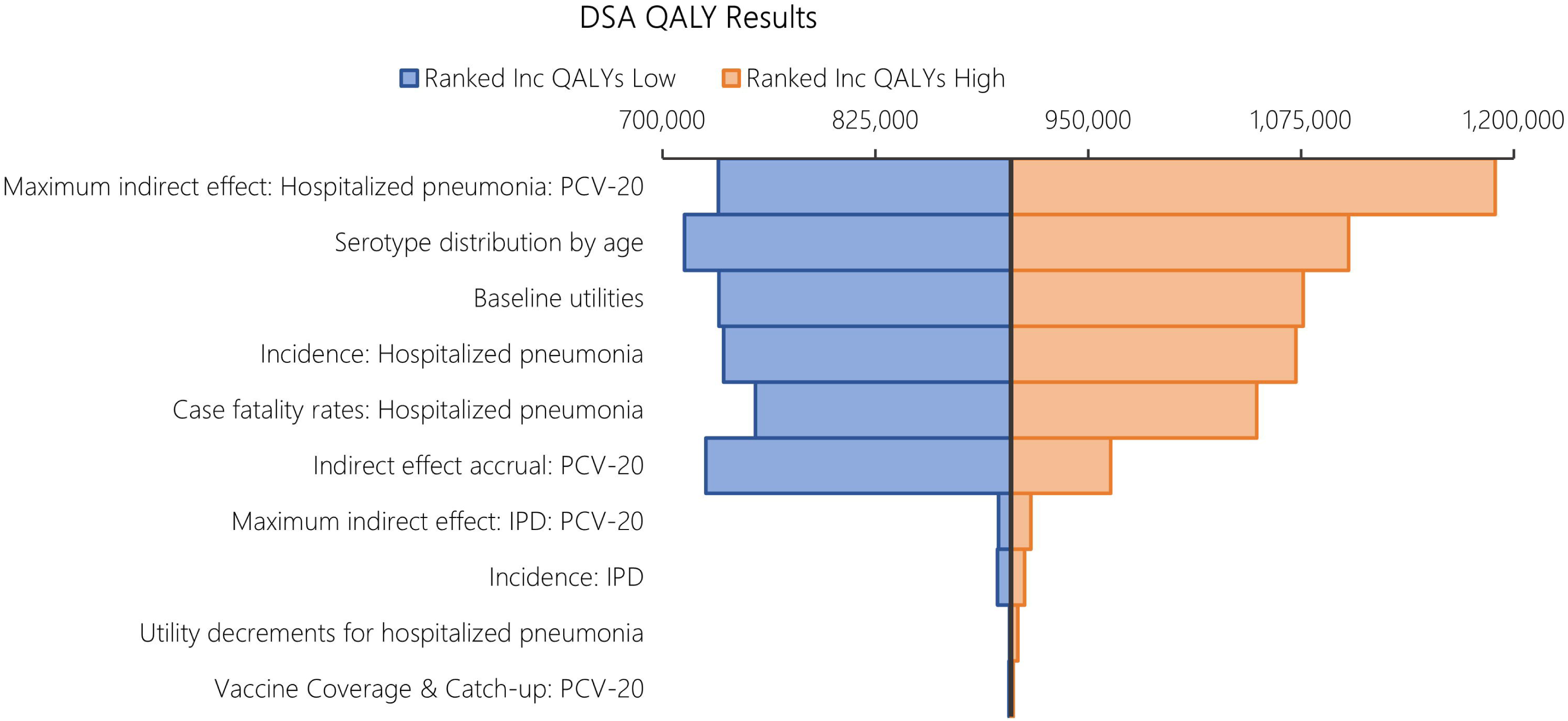
PSA cost-effectiveness plane vs baseline of PCV13. Abbreviations: PCV13, 13-valent pneumococcal conjugate vaccine; PCV20, 20-valent pneumococcal conjugate vaccine; PSA, probabilistic sensitivity analysis

**Figure 11:**
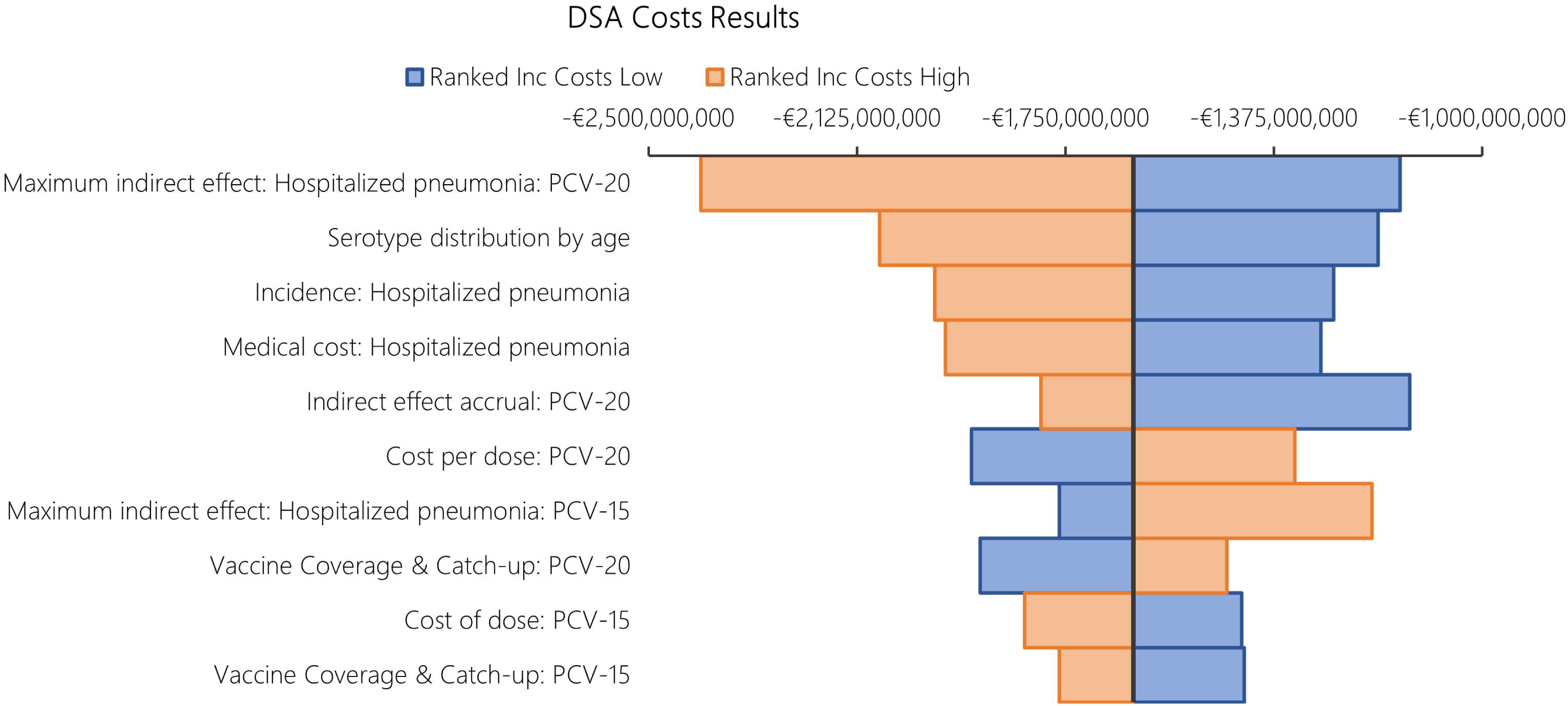
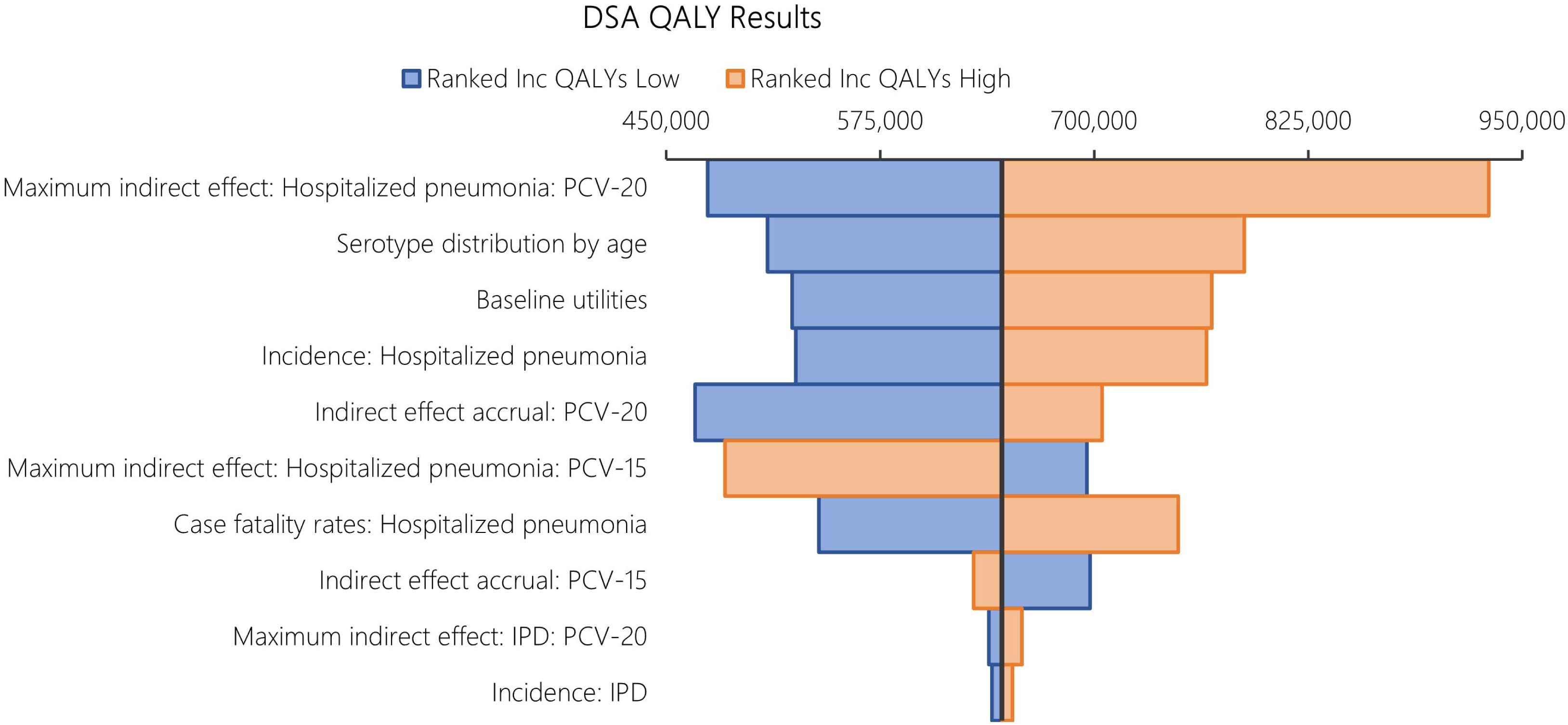
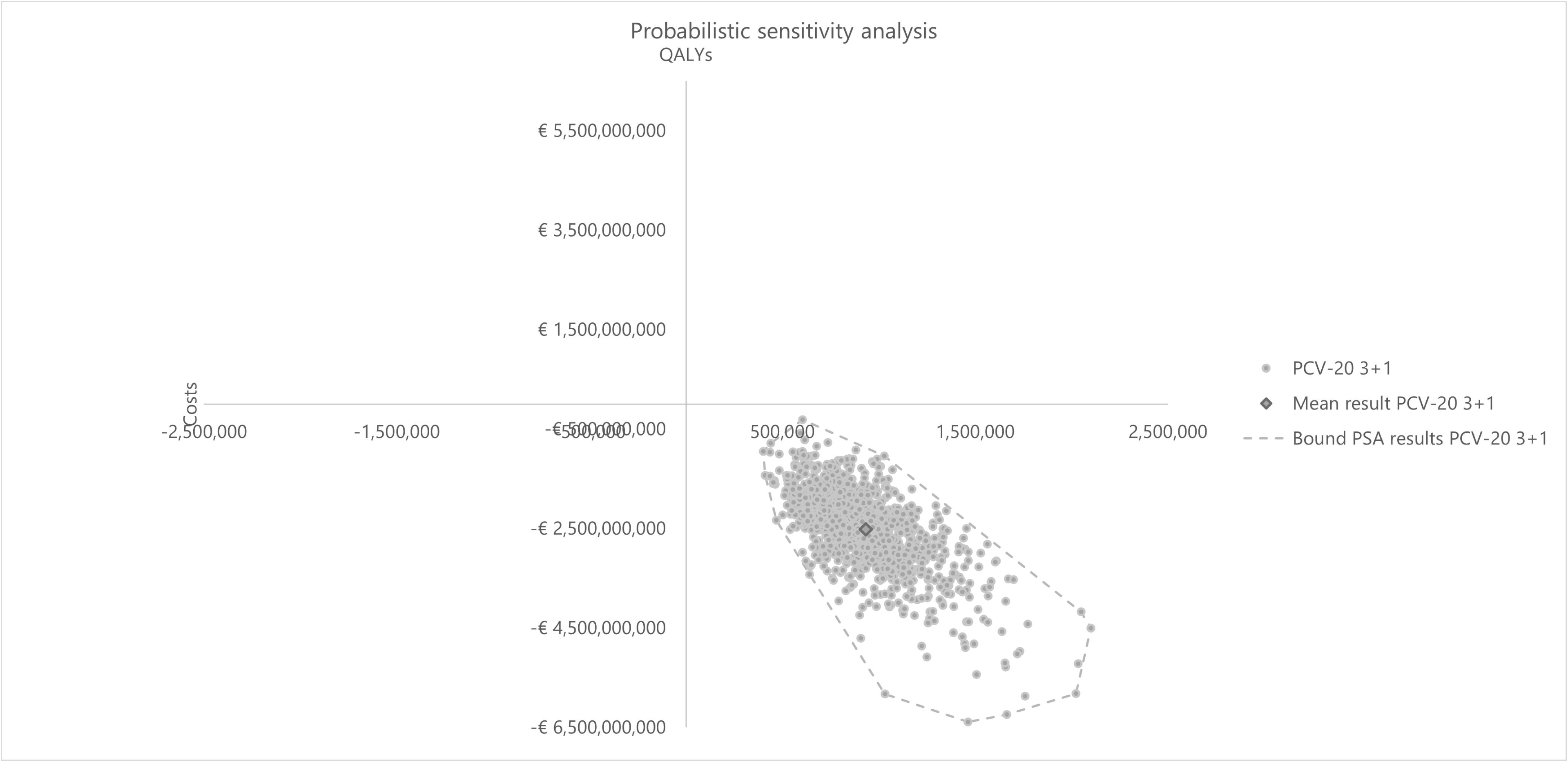
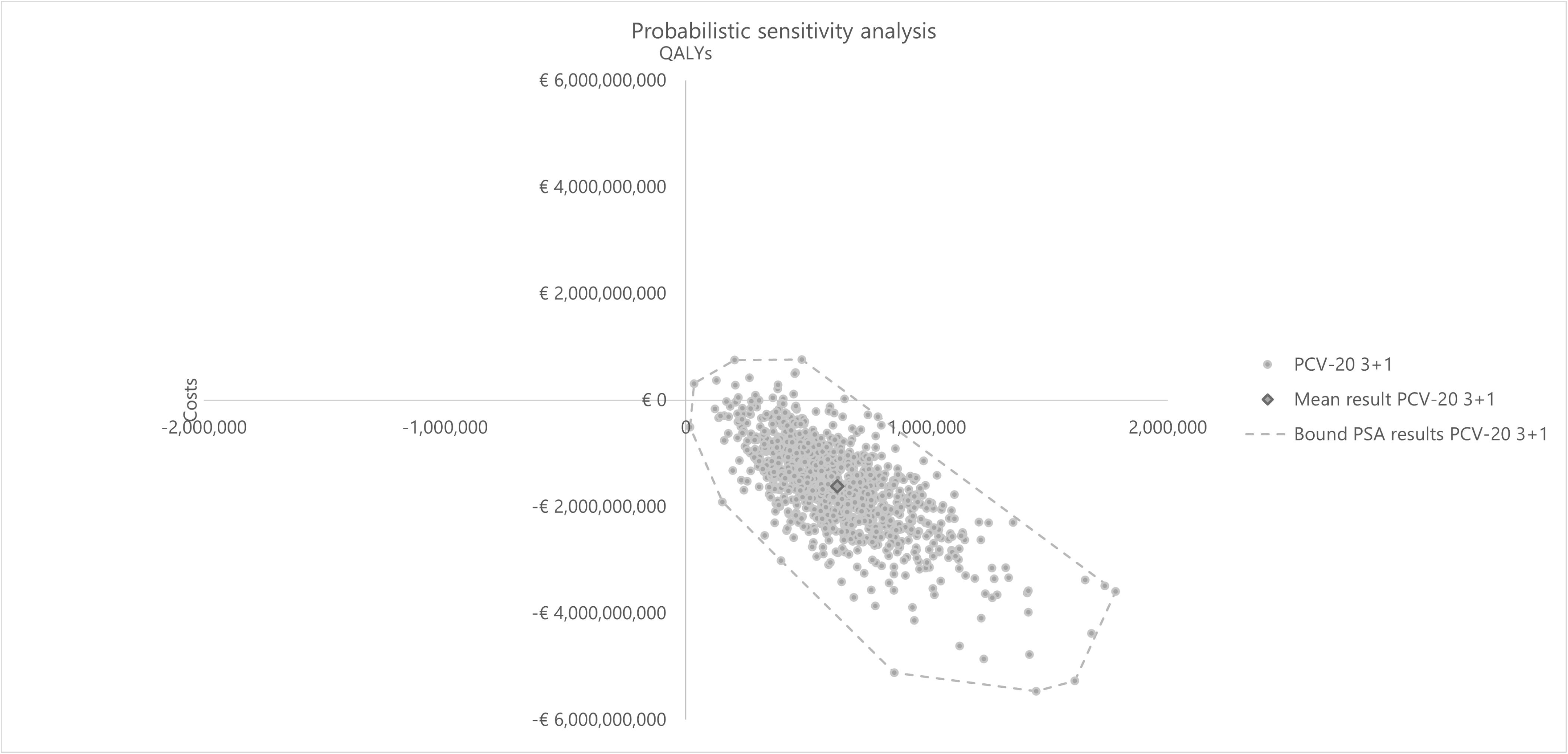
PSA cost-effectiveness plane vs baseline PCV15. Abbreviations: PCV13, 13-valent pneumococcal conjugate vaccine; PCV20, 20-valent pneumococcal conjugate vaccine; PSA, probabilistic sensitivity analysis

**Table 5:** PSA results.

|  | PCV20 vs PCV13 |  | PCV20 vs PCV15 |  |
| --- | --- | --- | --- | --- |
|  | QALY | Costs | QALY | Costs |
| **Base values** | 904,854 | -€2,393,263,611.36 | 646,235 | -€1,628,000,506.17 |
| **Mean values** | 933,044 | -€2,517,712,051.73 | 627,739 | -€1,618,779,090.22 |
| **SD values** | 253,634 | €828,848,311.25 | 244,826 | €842,934,915.67 |
| **2.5 percentile values** | 1,519,557 | -€1,114,095,339.11 | 1,158,046 | -€117,464,996.99 |
| **97.5 percentile values** | 548,941 | -€4,373,904,479.43 | 229,669 | -€3,410,047,589.50 |
| **More costly/more effective** | 0.00% |  | 1.60% |  |
| **More costly/less effective** | 0.00% |  | 0.00% |  |
| **Less costly/less effective** | 0.00% |  | 0.00% |  |
| **Less costly/more effective** | 100.00% |  | 98.40% |  |

Abbreviations: PCV, pneumococcal conjugate vaccine; QALY, quality-adjusted life year; SD, standard deviation; SoC, standard of care

### Scenario analyses

Scenario analysis results were summarized in **Table 6**. In the three scenarios where different discount rates were applied for costs and benefits, the qualitive conclusion of PCV20 being the dominant strategy compared to both comparators remained robust. When reducing indirect effects (i.e. maximum reduction in disease incidence) by half, PCV20 was still estimated to have better health benefits and lower costs compared to both PCV13 and PCV15. Similarly, extending time to realize indirect effects (i.e. accrual data in the first 2 years at 0%) and increased vaccination uptake in adults 65+ years old aligned with the base case results of PCV20 being the dominant strategy versus both comparators. Using a payer perspective led to a decrease by 15% and 17% in ICER in the comparison between PCV20 versus PCV13 and PCV20 versus PCV15, respectively. However, the qualitative conclusion remained the same. Other scenarios testing a different waning assumption (i.e. reducing duration of full protection to 3 years) and serotype unmasking were in line with the base case with minimal change in ICER. Overall, the results and conclusion were relatively robust. When assuming a high vaccine uptake in infant (90%), ICERs increased slightly, at less than 1%, for both PCV20 vs PCV13 and PCV20 vs PCV15. Considering disutility related to adverse events related to the administration of all vaccines (e.g. local reaction and systematic reaction or fever) resulted in the same conclusion of PCV20 3+1 being the dominant strategy among 3 PCVs.

**Table 6:** Scenario analyses.

| # | Description | Incremental costs | Incremental QALYs | Incremental costs | Incremental QALYs |
| --- | --- | --- | --- | --- | --- |
|  | PCV20 vs. | PCV13 |  | PCV15 |  |
|  | **Base case** | -€ 2,393,263,611.36 | 904,854 | -€ 1,628,000,506.17 | 646,235 |
| 1a | **Discount rate:** No discount rate (0%) for both costs and benefits | -€ 2,848,245,995.56 | 1,442,186 | -€ 1,943,755,486.67 | 1,037,762 |
| 1b | **Discount rate:** 0% discount in effects and 3% discount in costs | -€ 2,393,263,611.36 | 1,442,186 | -€ 1,628,000,506.17 | 1,037,762 |
| 1c | **Discount rate:** 1.5% discount in effects and 3% discount in costs | -€ 2,393,263,611.36 | 1,128,398 | -€ 1,628,000,506.17 | 808,398 |
| 2a | **Indirect effect:** Reduce indirect effects by half | -€ 963,573,932.75 | 437,813 | -€ 582,626,514.74 | 312,870 |
| 2b | **Indirect effect:** Extending time to realize indirect effect (i.e. accrual data in the first 2 years at 0%) | -€ 2,009,850,006.75 | 783,204 | -€ 1,346,292,762.25 | 559,090 |
| 2c | **Indirect effect:** increased vaccination rate among adults 65+ years old to 50% (assumption) | -€ 1,794,313,578.56 | 667,090 | -€ 1,215,807,895.27 | 482,623 |
| 3 | **Waning:** Reducing duration of full protection to 3 years (waning by year 8) | -€ 2,394,554,186.87 | 905,049 | -€ 1,628,963,569.45 | 646,382 |
| 4a | **ST unmasking** (ST distribution reduces by 5% annually compared to baseline up to year 5 - steady state) for PCV20 and PCV15 | -€ 1,869,690,557.98 | 741,193 | -€ 1,242,719,695.87 | 529,409 |
| 4b | **ST unmasking** (ST distribution reduces by 10% annually compared to baseline up to year 5 - steady state) for PCV20 and PCV15 | -€ 1,346,232,733.84 | 577,569 | -€ 857,543,262.30 | 412,616 |
| 4c | **ST unmasking** (% reduction in ST distribution annually from RWE) for PCV20 and PCV15 | -€ 2,243,987,841.94 | 880,662 | -€ 1,511,105,908.94 | 628,648 |
| 5 | **Payer perspective** | -€ 2,035,127,528.02 | 904,854 | -€ 1,343,839,409.40 | 646,235 |
| 6 | **Vaccination rates in infants:** increase to 90% (assumption) | -€ 2,402,252,440.51 | 905,071 | -€ 1,634,550,212.64 | 646,417 |
| 7 | **AEs related to vaccination:** Considering disutility related to the administration of all vaccines | -€ 2,393,263,611.36 | 904,052 | -€ 1,628,000,506.17 | 645,432 |

Abbreviations: AEs, adverse events; ICER, incremental cost-effectiveness ratio; PCV, pneumococcal conjugate vaccine; QALY, quality-adjusted life year; RWE, real-world evidence; ST, serotypes

This article is based on previously conducted studies or collected published data and does not contain any new studies with human participants or animals performed by any of the authors.

## Discussion

The introduction of next-generation PCVs with increased serotype coverage in Europe has provided options to be considered in national childhood immunization programs. This study examined the cost and health outcomes related to PCV20 under a 3+1 schedule compared with PCV13 and PCV15, both under a 2+1 schedule, in the pediatric population in Germany.

The base-case results demonstrated PCV20 as the dominant strategy over both lower-valent alternative vaccines. PCV20 was estimated to have greater health benefits than both PCV13 and PCV15, by averting total more cases of pneumococcal diseases including IPD, pneumonia and OM over a 10-year time horizon. This resulted in higher QALY gained and lower total costs related to PCV20, implying dominance of PCV20 compared with PCV13 and PCV15. Several scenarios assessed additional uncertainty, such as effects of different discount rates for costs and outcomes, several assumptions on vaccine effects (i.e. reduction in indirect effects and extension in accrual time of indirect effects), and waning duration. In addition, serotype replacement assumptions were examined to test how sensitive the results were to reduction in vaccine-type serotype coverage over time. Finally, payer perspective and an assumption of higher vaccine uptake in infant were explored. The results were robust across all sensitivity analyses including PSA and DSA.

Our findings are consistent with published studies comparing PCV20 to PCV13 and PCV15 in other settings. In Canada [37] and Greece [60], PCV20 was estimated to be cost-saving compared with PCV15 in a 2+1 schedule. In the United Kingdom (UK), PCV20 2+1 was estimated to be cost-saving compared to PCV13 1+1 and cost-effective compared with PCV20 1+1 [61]. Moreover, a public health impact analysis in the Netherlands and estimated that PCV20 could avert 45,127 pneumococcal cases compared to PCV10 over 5 years [62].

Our analyses have several limitations. Firstly, a Markov cohort model was considered appropriate for the decision problem based on previous cost-effectiveness assessments of PCVs [63]. Static models are commonly used for economic evaluations of PCVs in Germany [57, 64] and globally [42, 65–69]. While dynamic models are known to capture indirect effects, the decision-analytic Markov cohort model in this study utilizes the simplistic static Markov framework and incorporates components such as indirect effects. This notable improvement in the modelling approach helps quantify the far-reaching effects of vaccination at the population level while maintaining the clarity and transparency of the model.

German data were prioritized to parameterize the model. When German data were not available, data were sourced from other high-income European countries with similar health care systems. Direct effects were estimated from different sources using PCV13 and PCV7 studies, given no studies have measured the effectiveness of PCV15 or PCV20 against pneumococcal disease outcomes. Differential herd effects were not modelled based on PCV schedule but have been observed in countries that have implemented PCV13 in infant NIPs (i.e., increase in disease reduction under a 3+1 vs 2+1)[25, 49]. There are potential confounding factors, such as the rate of reduction and time to stabilization of IPD incidence across age groups and countries may be associated with multiple factors including vaccine uptake, implementation of a catch-up program, duration of PCV use, availability of an adult a pneumococcal vaccination program, serotypes in circulation, and general epidemiologic variability. To assess the uncertainty around the indirect effect estimations for IPD and non-invasive disease, extensive sensitivity analyses were conducted, such as PSA, DSA, and several scenarios.

The base case analysis did not include serotype replacement. It is difficult to predict how the characteristics or composition of non-vaccine serotypes will change following higher-valent PCV introduction. Model simulations suggest that replacement may be less for high-valency PCVs [70, 71]. To address uncertainty, we tested the impact of increasing non-vaccine serotypes overtime, which all led to similar directional results as the base case.

We did not consider adverse events originating from vaccination by PCV20, PCV15, or PCV13 in our base case analysis. Though, adverse events after pneumococcal vaccination resulting in healthcare seeking are rare and event rates are similar for the different pneumococcal vaccines, PCV20, licensed in a 3+1 schedule, is likely to result in more adverse events than PCV15 and PCV13, both under a 2+1 schedule. We tested this scenario and found that although PCV20 3+1 was estimated with slightly higher disutility related to the extra dose, the strategy still provides higher total QALY gain compared to lower-valent alternatives and remained dominant.

Despite the higher immunological response against serotype 3 of PCV15, we did not model higher vaccine effectiveness, as data on clinical effectiveness of PCV15 against serotype 3 are unknown. In contrast, a meta-analysis of observational studies supports direct PCV13 protection against serotype 3 IPD in children. Without any real-world effectiveness data for PCV15, there is no way to assess the actual impact of PCV15 on ST3. For that reason, PCV15 and PCV20 are assumed to provide comparable protection as PCV13 against disease caused by serotype 3.

Recently, STIKO recommended PCV20 for adults aged 60 years and older and for adult patients with underlying diseases[18]. Vaccination rates among adults may increase in the future as PCV20 is only administered once in adults. To account for the direct impact of adult vaccination, we assumed a proportion adults received a PCV and therefore received no additional benefit of indirect effects from the pediatric program. We also tested the impact of less pronounced herd effects associated with higher-valent PCVs. Changes to the assumptions resulted in fewer cases avoided and smaller life expectancy gains under a PCV20 pediatric program. However, PCV20 under a 3+1 program remained the dominant strategy avoiding more cases, increasing life-expectancy while costing less than PCV13 or PCV15 under a 2+1 program.

Indirect cost estimates were not based on a rigorous German database cost assessment. We applied estimates based on published assumptions. To avoid assumptions on the indirect costs, we carried out a scenario from the payer perspective, not accounting for any indirect costs. This is a very conservative approach given that parents stay at home or have another caregiver for their child. Even under conservative assumptions, PCV20 vaccination strategy was clearly dominating PCV15 and PCV13 strategies resulting in fewer cases and fewer costs.

## Conclusion

The results of this CEA estimated that the implementation of PCV20 under a 3+1 schedule into German immunization recommendation would be less costly, and more effective than PCV13 and PCV15, both under 2+1 schedule. PCV20 has the potential to substantially decrease the clinical and economic burden of pneumococcal diseases in Germany by providing substantially broader protection compared with lower-valent vaccines.

## Acknowledgements

The original model was developed by Des Dillon-Murphy, PhD, and Ruth Chapman, PhD, of Evidera. Writing support for this manuscript was provided by Colleen Dumont and Sally Neath of Cytel Inc, Waltham, MA, US and by Allison Brackley, PhD, and Elizabeth Hubscher, PhD, who were employed by Cytel Inc, Waltham, MA, US at the time of the manuscript development. We thank Mark van der Linden for providing IPD data.

## Author Contributions

Felicitas Kühne, Maren Laurenz, Christof von Eiff, Johnna Perdrizet and Sophie Warren contributed to the study conceptualization, investigation, and methodology. An Ta conducted the formal analysis and was responsible for project administration. Johnna Perdrizet was responsible for funding acquisition. Johnna Perdrizet and Felicitas Kühne provided supervision and validation. All authors contributed to writing the original draft, as well as reviewing and editing subsequent drafts, and approving the final version.

## Ethics/Ethical Approval

This article is based on previously conducted studies and does not contain any new studies with human participants or animals performed by any of the authors; as such, ethical approval was not required.

## Data Availability

All data generated or analyzed during this study are included in this published article/as supplementary information files.

