## Supplement for "Cost-effectiveness of PCV20 to Prevent Pneumococcal Disease in the Pediatric Population - A German Societal Perspective Analysis"

**Prior presentation:** An abstract reporting the results of this study has been submitted to ESPID 2024 (20–24 May), which has been accepted as a poster presentation.

Appendix A: Indirect Effects

The estimates of indirect effects were based on (i) accrual data capturing the ramp-up period until the steady state is reached (i.e. no additional indirect effects would be observed after this state), and (ii) the maximum reduction in pneumococcal disease incidence.

(i)	Accrual data

The model assumed that indirect effects on current PCV13 serotypes are in a steady state since PCV13 has been used for more than 10 years. In other words, no additional indirect effects related to PCV13-unique serotypes would be observed with higher-valent vaccines: PCV15 or PCV20. The indirect effects from PCV15 and PCV20, therefore, were only quantified from the newly covered serotypes and assumed to occur gradually over the model time horizon.

The accrual of the indirect effects for PCV15 and PCV20 were informed by IPD surveillance data obtained in the UK (Ladhani et al., 2018[1]) with year 6 of the PCV13 infant program chosen as the steady-state year per Perdrizet et al., 2023[2]. It is noted that Serotype 3 was excluded from the analysis due to its unique immunologic characteristics compared with other PCV13-serotypes and its persistent pneumococcal carriage in pediatrics [3-5], resulting in the use of annual relative change in vaccine-type IPD incidence within the serotype group PCV13 minus PCV7 serotypes (excluding serotype 3 [PCV13-7-ST3]) after the introduction of PCV13. Consequently, the potential decrease in the incidence of the newly covered serotypes in PCV15 and PCV20 was estimated.

(ii)	The maximum reduction in pneumococcal disease incidence

The maximum of herd effects against IPD caused by additional serotypes covered by PCV15 and PCV20 was informed by Ladhani et al., 2018[1] and Perdrizet et al., 2023[2].

For hospitalized pneumonia, the effect on children was estimated using an observational study by Levy et al., 2017 that was conducted in France in 2009 – 2015 [6]. The study recorded a 30% decrease in inpatient hospitalizations from emergency room visits following PCV13 implementation in children aged 5-17 years (i.e., post-PCV13 implementation data in 2011 – 2015 compared with the pre-PCV13 period in 2009 – 2010). The model applied adjustment ratios of 70% and 86% to account for differences in serotype distribution pre- PCV13 implementation and the current serotype distribution (i.e. PCV13 serotype coverage was 70% for infants aged <2 and 86% for children 2 to 17 years of age in France [7]). For age groups ≥18 years old, data from a UK-based study for adults was used. The study reported a 15% reduction in the incidence of chest X-ray-confirmed pneumonia cases after the implementation of PCV13 in pediatric vaccination[8]. Likewise, ratios of 67% (18-49 years), 60% (50-65 years), and 56% (≥65 years) were used for adjustment[1]. A similar approach with the same data sources were used to estimate maximum reduction non-hospitalized pneumonia, however, only for children.

For otitis media (OM), it was assumed that indirect effects were only observed in children younger than 5 years old. Maximum decrease in OM incidence was informed by a retrospective cohort study by Lau et al., 2015[9], which included children aged 5 to 9 years in the UK (i.e. comparing OM incidence from before the introduction of PCV13 in 2007 – 2010 with post-PCV13 implementation in 2011 – 2012). To adjust serotype coverage, a ratio adjustment of 71% in 2009, as reported by Ladhani et al., 2018, was used [1].

The model excluded the vaccinated adult population from indirect effects to avoid overestimating indirect effect from the infant vaccination program.

Appendix B: Supplementary tables

Table S1: General population estimates

| Age groups | Population size |
| --- | --- |
| <12 months | 791,254 |
| 12–23 months | 780,795 |
| 24–35 months | 789,145 |
| 36–47 months | 803,334 |
| 48–59 months | 810,805 |
| 5–17 years | 9,887,926 |
| 18–34 years | 16,539,516 |
| 35–49 years | 15,331,897 |
| 50–64 years | 19,065,953 |
| ≥65 years | 18,972,098 |
| **Total** | **83,772,723** |

Source: German population data from German Federal Statistical Office [10]

Table S2: Number of <12-month-olds in incoming birth cohort

| Year in model | Number of infants younger than 12 months in incoming birth cohort |
| --- | --- |
| Year 2 | 747,800 |
| Year 3 | 752,300 |
| Year 4 | 750,200 |
| Year 5 | 747,300 |
| Year 6 | 743,400 |
| Year 7 | 738,900 |
| Year 8 | 734,100 |
| Year 9 | 729,000 |
| Year 10 | 723,600 |

Source: German Federal Statistical Office [10] and German Federal Statistical Office [11]

A birth rate of 1.4 per female was assumed to calculate the number of infants younger than 12 months in incoming birth cohort for per year, based on German Federal Statistical Office [11] estimates.

Table S3: Probability of sequela

| **Age group** | **Meningitis** [12] | |
| --- | --- | --- |
|  | **Deafness** | **Non-deafness** |
| All age groups | 9.12% | 12.48% |

Table S4: Age-specific Clinical Outcome Results: Total disease cases and deaths

| **Age** | **PCV13 2+1** | | | | |
| --- | --- | --- | --- | --- | --- |
|  | **IPD** | **Hospitalized pneumonia** | **Non-hospitalized pneumonia** | **OM** | **Total deaths due to disease** |
| <12 months | 1,177 | 111,173 | 182,210 | 1,098,249 | 200 |
| 12-23 months | 1,184 | 60,995 | 183,360 | 1,105,180 | 112 |
| 24-35 months | 106 | 61,494 | 641,527 | 1,355,200 | 59 |
| 36-47 months | 107 | 62,070 | 647,544 | 1,367,909 | 60 |
| 48-59 months | 108 | 62,670 | 653,801 | 292,101 | 60 |
| 5 - 17 years | 1,025 | 132,665 | 2,434,233 | - | 1,249 |
| 18 - 34 years | 2,537 | 167,228 | 602,326 | - | 7,441 |
| 35 - 49 years | 2,662 | 175,509 | 632,155 | - | 7,807 |
| 50 - 64 years | 18,734 | 955,072 | 1,226,979 | - | 114,373 |
| 65+ years | 49,374 | 5,723,352 | 2,294,427 | - | 981,142 |
| **Total** | **77,013** | **7,512,228** | **9,498,563** | **5,218,640** | **1,112,503** |

| **Age** | **PCV15 2+1** | | | | |
| --- | --- | --- | --- | --- | --- |
|  | **IPD** | **Hospitalized pneumonia** | **Non-hospitalized pneumonia** | **OM** | **Total deaths due to disease** |
| <12 months | 1,124 | 108,506 | 179,647 | 1,083,972 | 194 |
| 12-23 months | 1,131 | 59,479 | 180,736 | 1,090,433 | 108 |
| 24-35 months | 98 | 59,136 | 627,168 | 1,327,219 | 57 |
| 36-47 months | 100 | 59,829 | 633,389 | 1,340,573 | 58 |
| 48-59 months | 102 | 60,545 | 639,840 | 286,452 | 58 |
| 5 - 17 years | 947 | 128,341 | 2,377,912 | - | 1,206 |
| 18 - 34 years | 2,400 | 165,368 | 602,328 | - | 7,349 |
| 35 - 49 years | 2,514 | 173,495 | 632,157 | - | 7,707 |
| 50 - 64 years | 17,674 | 940,185 | 1,227,019 | - | 112,574 |
| 65+ years | 46,955 | 5,633,084 | 2,295,013 | - | 965,523 |
| **Total** | **73,046** | **7,387,967** | **9,395,209** | **5,128,648** | **1,094,834** |

| **Age** | **PCV20 3+1** | | | | |
| --- | --- | --- | --- | --- | --- |
|  | **IPD** | **Hospitalized pneumonia** | **Non-hospitalized pneumonia** | **OM** | **Total deaths due to disease** |
| <12 months | 771 | 91,523 | 163,862 | 994,998 | 159 |
| 12-23 months | 795 | 50,380 | 164,995 | 1,001,950 | 84 |
| 24-35 months | 65 | 48,921 | 564,945 | 1,205,969 | 46 |
| 36-47 months | 69 | 50,119 | 572,054 | 1,222,117 | 48 |
| 48-59 months | 74 | 51,336 | 579,341 | 261,972 | 49 |
| 5 - 17 years | 746 | 116,449 | 2,223,028 | - | 1,087 |
| 18 - 34 years | 1,663 | 155,368 | 602,334 | - | 6,855 |
| 35 - 49 years | 1,717 | 162,668 | 632,165 | - | 7,173 |
| 50 - 64 years | 14,192 | 891,263 | 1,227,153 | - | 106,660 |
| 65+ years | 41,620 | 5,434,004 | 2,296,320 | - | 931,078 |
| **Total** | **61,712** | **7,052,031** | **9,026,198** | **4,687,006** | **1,053,238** |

**Note:** The breakdown results by age groups were mostly consistent with the overall results where more cases avoided related to PCV20 in all included age groups for IPD, hospitalized pneumonia and OM than PCV13, especially in the oldest group of 65+ year-olds. Similarly, PCV20 is estimated to prevent more cases of non-hospitalized pneumonia than PCV13 in children less than 5 years old. The higher valent vaccines showed an increasing number of prevented non-invasive pneumonia. One could observe a shift from severe cases (i.e., hospitalized cases) in the lower valent vaccine strategies to less severe cases (i.e.; non-hospitalized pneumonia) in the higher valent vaccine strategies. Furthermore, better health outcomes from PCV20 were shown by a reduction in deaths due to disease in all age groups, with the greatest reduction observed in those 65 years old and above (50,064 deaths averted).

10.	Statistisches Bundesamt (Destatis), *Population: Germany Reporting date, age years.* 2022.

11.	Destatis Statistiches, B. *Current population of Germany*. 2023 13 June 2023]; Available from: <https://www.destatis.de/EN/Themes/Society-Environment/Population/Current-Population/_node.html>.

12.	Kuhlmann, A. and J.M.G. von der Schulenburg, *Modeling the cost-effectiveness of infant vaccination with pneumococcal conjugate vaccines in Germany.* The European Journal of Health Economics, 2017. **18**(3): p. 273-292.
